# SARS-COV-2 Delta and Omicron community transmission networks

**DOI:** 10.1101/2022.05.30.22275787

**Authors:** John M. Murray, Daniel D. Murray, Evelyne Schvoerer, Elma H. Akand

## Abstract

To date, calculations of SARS-CoV-2 transmission networks at a population level have not been performed. Networks that estimate infections between individuals and whether this results in a mutation, can evaluate fitness of a mutational clone by how much it expands in number as well as determining the likelihood a transmission results in a new variant.

Transmission networks of SARS-CoV-2 infection between individuals in Australia were estimated for Delta and Omicron variants using a novel method. Many of the sequences were identical, with clone sizes following power law distributions driven by negative binomial probability distributions for both the number of infections per individual and the number of mutations per transmission (mean 1.0 nucleotide change for Delta and 0.79 for Omicron). Using these distributions, an agent based model was able to replicate the observed clonal network structure, providing a basis for more detailed COVID-19 modelling. Recombination events, tracked by insertion/deletion (indel) patterns, occurred for each variant in these outbreaks. The residue at position 142 in the S open reading frame (ORF), frequently changed between G and D for Delta sequences, but this was independent of other mutations. On the other hand, several Omicron mutations were significantly connected across different ORF. This model reveals key transmission characteristics of SARS-CoV-2 and may complement traditional contact tracing and other public health strategies. This methodology can also be applied to other diseases as genetic sequencing of viruses becomes more commonplace.

**Author summary:** As SARS-COV-2 spreads through a community, it can mutate and generate new variants. How likely this is to occur and how much a particular viral clone expands, can indicate mutational probabilities and whether some mutations are fitter than others. By better understanding these aspects, future predictions can more accurately encapsulate possible changes in the epidemic within a community. We have developed a new method for piecing together the individual SARS-COV-2 cases that have been sequenced, to generate the structure of transmissions and mutational clones for an outbreak. While this method can be applied to other virus epidemics given sufficient sequenced data, we apply it to Delta and Omicron outbreaks in Australia. Interestingly, transmissions between individuals frequently do not result in mutations, with some clones growing very large. We characterise the probability that a mutation will occur, and track how these changes lead to sequential mutations in these outbreaks.

## Introduction

While contact tracing has provided some detail of who transmits SARS-CoV-2 to whom, it has not been able to extensively determine community-wide transmission networks. Currently, studies exploring transmission networks for SARS-CoV-2 are limited to a few hundred cases and require extensive epidemiological and genetic investigation [1, 2]. While genomic and epidemiological methods can be expanded to cluster more samples, they are limited in their ability to track pathways of transmissions [3, 4]. However, since the pandemic started, many countries have scaled up genetic sequencing of individuals infected with SARS-CoV-2 and population level assessment of transmission networks is now possible. Individual samples in a community with similar sequences, reported at similar times, are likely connected through a transmission pathway. Determining the most probable among all of these possible pathways can provide an estimate of the transmission network that spreads SARS-CoV-2 through a community.

Australia provides a unique opportunity to study SARS-CoV-2 transmission networks. Until more recently, authorities within Australia sequenced a high percentage of confirmed infections [3, 5], while strict public health policies, including mandatory quarantine for those coming into the country from March 2021, prevented frequent cross-border infection sources. This means that any transmissions within the country generally stayed in the country and were recorded there. Therefore, with an aim to calculate transmission networks, SARS-CoV-2 sequences were downloaded from GISAID (the Global Initiative on Sharing All Influenza Data), for two major outbreaks within Australia.

The method to determine the network transmission structure for these outbreaks is first developed. As well as providing mutational pathways for the virus evolution, these calculations also reveal the clonal structure for these outbreaks and the probability a transmission results in a mutation. The underlying probability distributions are then used in an agent-based model to generate *in silico* outbreaks reflecting these characteristics. The mutations that appear across several transmission groups are analysed for their likelihood to occur in the presence or absence of others. This extracts compensatory mutations across several open reading frames (ORF).

## Results

Delta sequences up until 31 October 2021 (18,383 sequences), and Omicron sequences up until 14 January 2022 (5,436 sequences), Fig 1). Except for 6 BA.2 and 4 B.1.1.529 sequences, all Omicron variants belonged to the BA.1 lineage. The Delta variants were described by 86 lineages AY.*, B.1.617.2, but after removal of small groups were restricted to AY.23, AY.39, AY.39.1, AY.39.1.1, AY.39.1.2, AY.39.1.3, and B.1.617.2.

**Fig 1:**
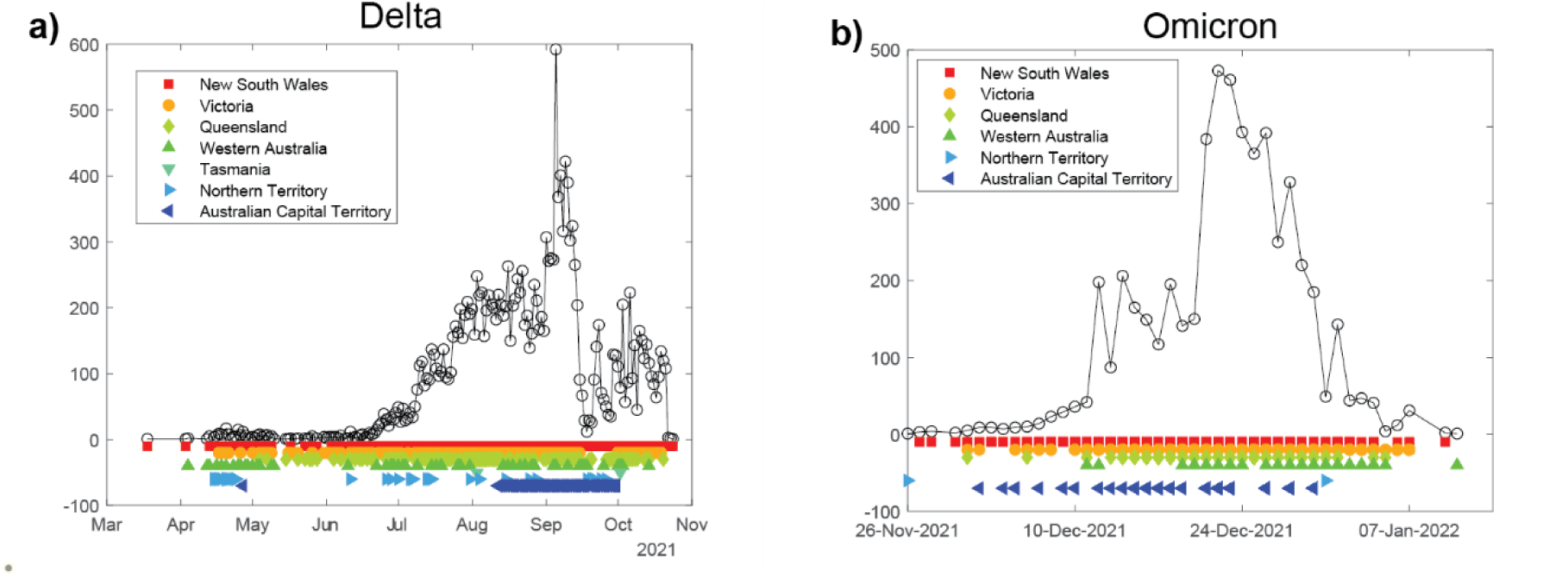
total number of sequenced cases for a) Delta (up to 31 October 2021), and b) Omicron cases (up to 14 January 2022) in Australia. Delta cases were limited to that time point in order to reduce computational complexity of alignment and transmission calculations. A marker below the plot denotes a sequence within the total number was located in the specified state at that time. Decreases in numbers of reported Delta cases are due to the percentage of sequenced cases dropping from 90% in July 2021 to less than 20% in early September, and the censoring of Omicron data to those uploaded by 14 January 2022. Sequencing in late 2021 was also limited to cases where “it will inform treatment choices as some therapies work with Delta but not for Omicron, and in situations where it will inform public health action” (NSW Health).

### Optimal arborescence

The likelihood the virus was transmitted between two individuals whose viral sequences had been uploaded, was estimated from the time when each sample was submitted for sequencing, and the number of differences between the nucleotide (nt) sequences over the regions coding for the ORF 1a, 1b, S, 3a, E, M, 6, 7a, 7b, 8, N and 10. For a given pair of sequences *s*_*1*_, *s*_*2*_ with sampling dates *t*_*1*_, *t*_*2*_, the interval between dates was taken as a surrogate for the time between symptoms (serial interval), which has been estimated to follow a Gamma distribution [6]. The probability of transmission, from identical sequences *s*_*1*_ to *s*_*2*_, was then given by *p(s*_*1*_, *s*_*2*_*)=Γ(t*_*2*_ *- t*_*1*_ *-10*.*5)*, where the time shift allowed transmissions from *s*_*1*_ to *s*_*2*_ even if the first was uploaded later due to delayed testing.

Over identical sequences *S=[s*_*1*_, *s*_*2*_, *…, s*_*n*_], and their possible directed transmissions *E=[(s*_*1*_, *s*_*2*_*), (s*_*1*_, *s*_*3*_*), …, (s*_*n-1*_, *s*_*n*_)], the maximal probability transmission network covers all sequences and is determined from

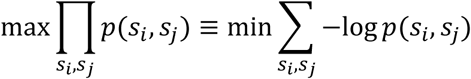

If samples can also differ in sequence by *d(s*_*1*_, *s*_*2*_), here taken as the number of nt differences, then the optimization problem is modified by a penalty term

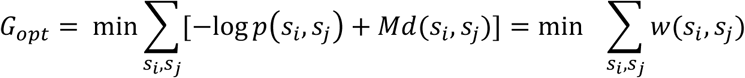

The solution to this problem is a minimal directed spanning tree, also called a minimal arborescence, and its calculation is achievable with a slightly more complex method than for the undirected situation [7, 8].

Within each set of lineages, sequences were initially pairwise aligned to the reference sequence EPI_ISL_412026 [9], after which the start and end of the 12 ORFs were estimated. Rather than perform the complex task of multiply aligning thousands of ∼29,000nt sequences, they were separated into groups determined by unique lengths over the 12 ORFs which were then used as alignments. Within each of these lineage and length groups, nt distances between pairs of sequences were estimated over common regions where both sequences were conclusively determined (not an ‘N’ for example). The number of possible edges in the original directed tree can grow combinatorially, so transmission networks were determined in two steps: firstly between different clones (groups of sequences that were identical over the 12 ORFs modulo any uncertain regions - anywhere between zero and several thousand positions were registered as ‘N’), and then within a clone.

Optimal clonal transmission networks were calculated for all groups containing at least 10 clones, resulting in 31 separate clonal transmission networks for Delta (containing 15,621 sequences, S1 Fig), and 23 for Omicron (containing 4,498 sequences, S2 Fig). Although the networks were determined separately, some of those networks would have been established by an insertion/deletion or mutations in the start or stop codons of an ORF giving rise to a group with different ORF lengths, and so there would be linkages between some of these networks (a node in one network to a root node in another). The alternative would be that a different group was initiated by a new variant from overseas.

The optimal arborescence for clones of the largest Delta group is shown in Fig 2, while that for the largest Omicron clonal group appears in Fig 3.

**Fig 2:**
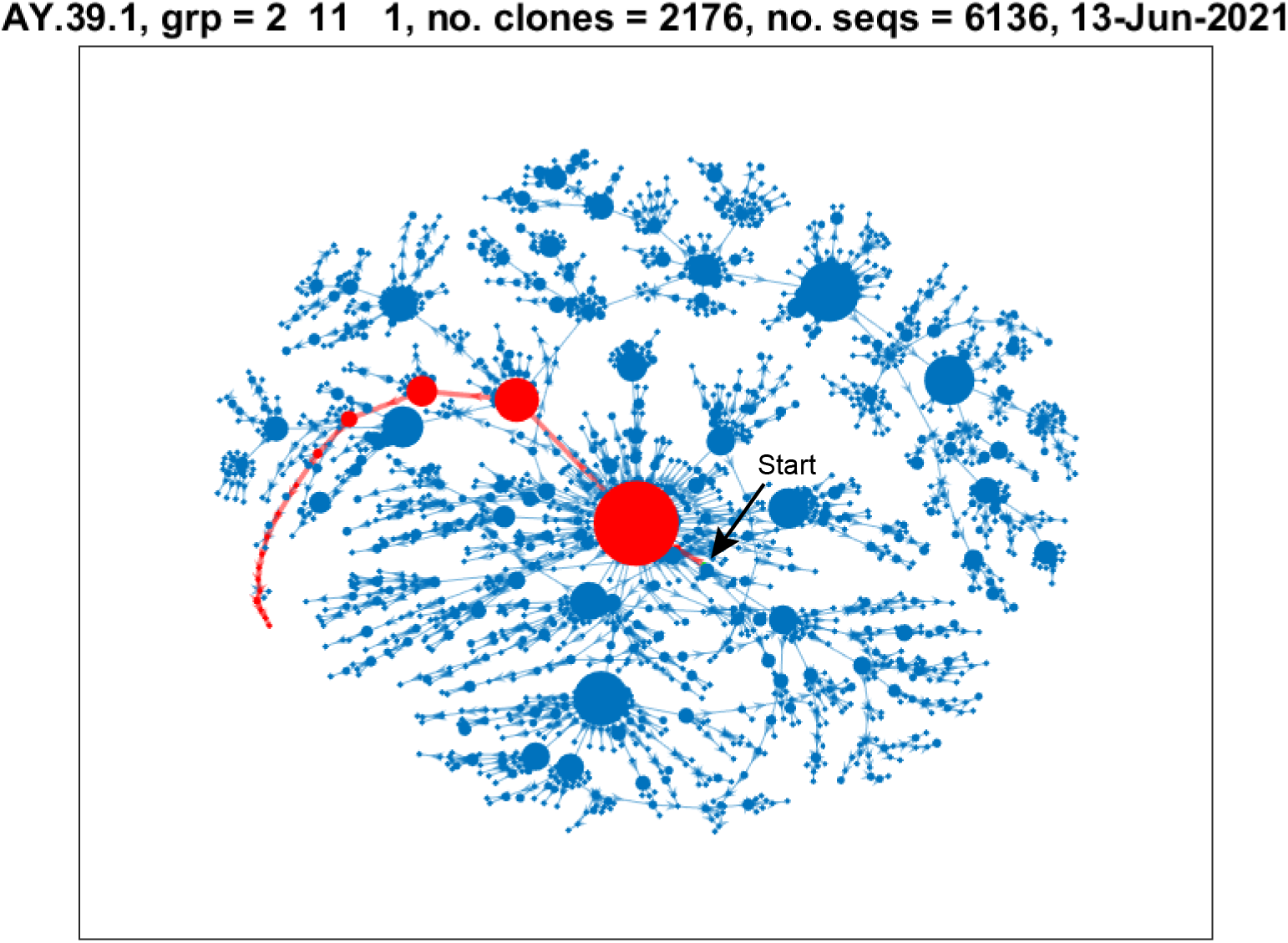
the optimal transmission network for the largest Delta group where the first sequence was sampled on 13 June 2021. The clone containing the start of the infection is shown as a green marker towards the centre of the figure. The size of the nodes represents the square root of the number of sequences within that clone. The red nodes and edges display the path with the most mutations starting from the root node).

**Fig 3:**
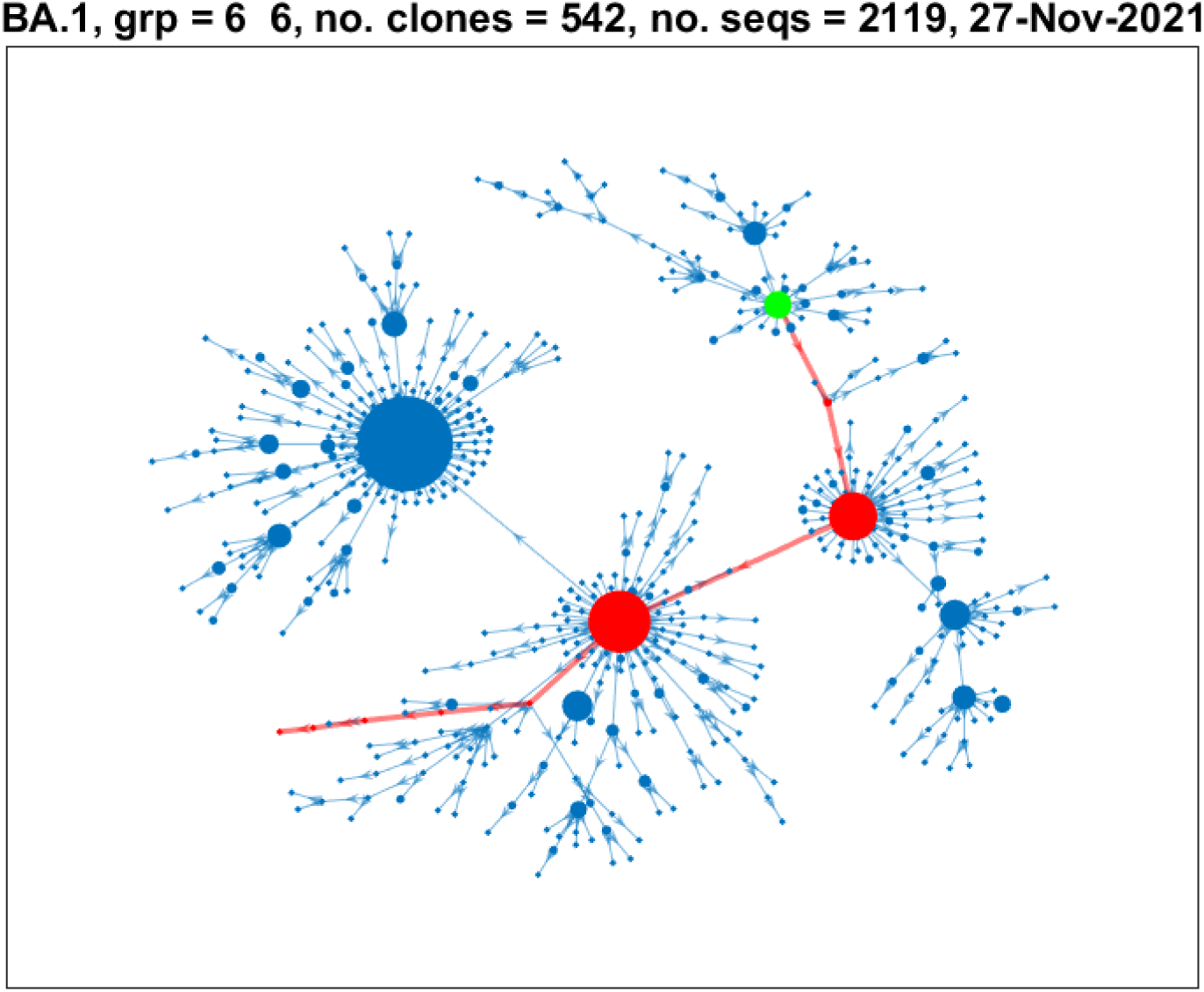
the optimal transmission network for the largest Omicron group where the first sequence was sampled on 27 November 2021. The clone containing the start of the infection is shown as a green marker. The size of the nodes represents the square root of the number of sequences within that clone. The red nodes and edges display the path with the most mutations starting from the root node.

### Power law distribution of clonal sizes

The size of some clones was surprisingly large, so that to reasonably represent them by node size in a figure required scaling by the square root of the number of sequences within the clone. Within the largest Delta group, a total of 552 of 6,136 sequences (9%) were within a single clone (Fig 2), and over all Delta groups the largest clone often exceeded 20%. Despite the lower percentage of sequencing with Omicron and the shorter duration over which they were collected, clones could also be large, containing 30% of sequences (630/2119, Fig 3). The clonal sizes within these networks displayed power law distributions, with the probability of clones of size at least *x* being given by *P(X ≥ x) ∼ bx*^*-α*^, exhibiting approximately linear decay on a log-log plot (S3 Fig). Exponents α for these groups were 1.0 and 0.74 respectively, showing a higher likelihood of sequences not mutating during transmission of Omicron than Delta, or at least for this to be recognised with uncertainties in the sequencing process, more limited timespan, and lower percentages of sequenced Omicron cases. Mean *α* values over all groups were similar for Delta and Omicron (1.22 and 1.25 resp. Supplementary Tables S1 and S2).

The number of mutations per transmission can be calculated from the mutational differences along the edges between clones, relative to the number of these edges plus the number of edges within the clones (this will be one less than the number of sequences). Over all 31 Delta subgroups the number of nt mutations per transmission ranged between 0.35 and 1.75, mean 1.00 (S1 Table). The number of mutations per Omicron transmission had a more extensive range, 0.31 to 2.58, although a smaller mean of 0.79 (S2 Table). The number of mutations per transmission were generally best represented by Negative Binomial probability distributions (S4 Fig and S5 Fig, S1 Table and S2 Table).

### Transmissions between individuals

The above networks display transmissions between clones. Transmissions within each clone are calculated in the same fashion. The optimality method assigned edges out of the clone according to the first of the sequences with a specified optimal integer-valued date. Since many sequences were sampled at the same date, edges tended to be assigned to the same node. To avoid these artificially produced high-degree nodes, the sampling dates within a clone were assumed to be lognormally delayed (*µ = 1, σ = 1*). Additionally any multiple edges from a node to outside the clone were randomly assigned among nodes with equivalent dates prior to lognormal modification. A representative transmission network for the largest clone in Fig 2 is shown in Fig 4a, with their node out-degrees once again best described by a negative binomial distribution (Fig 4b). Calculations for the largest Omicron clone in Fig 3 are shown in Fig 4c,d.

**Fig 4:**
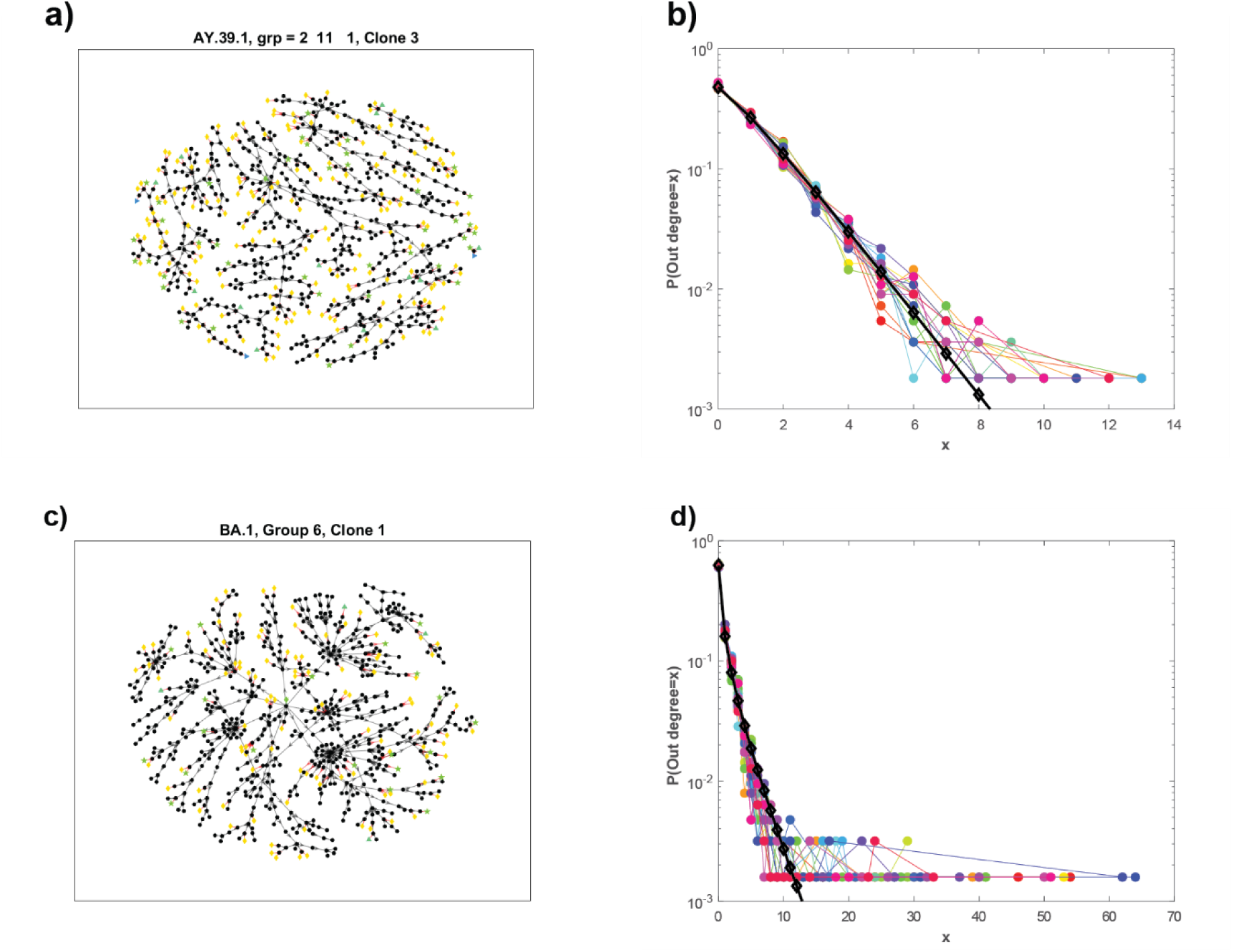
Representative optimal transmission network for the largest Delta clone in **Error! Reference source not found**. (a,b) and for the largest Omicron clone in Figure 3 (c,d). In a,c), sequences and transmissions within the clone are represented by black nodes and edges. Transmissions out of the clone containing mutations are shown with red edges and coloured markers (number of mutations: 1 (yellow diamond), 2 (green star), 3 (green up arrow), 4 (blue right arrow). In b,d,) out degree distributions over 20 simulations and the negative binomial distribution (b: R = 1.27, p=0.56, d: R = 0.34, p = 0.26), mean 1.0 (black lined and markers) fitted to the final simulation.

## A COVID network model

The fundamentals of the transmission network are the probability distributions for the number of new infections per infected person and the number of mutations per transmission, both of which are determined by negative binomial distributions. To determine how well these distributions produce the observed clonal structure, an agent-based model was constructed where at each generation the number of new transmissions for each agent was randomly drawn from a negative binomial distribution, as was the number of mutations occurring in that infection. The mean number of transmissions, or the basic reproduction number *R*_*0*_, for each individual in the largest network (Fig 2), was 1.0 (Fig 4). This low estimated value indicates that many infections in the community were undiagnosed and not all of the diagnosed cases were sequenced. To produce an ongoing outbreak, the parameters were modified to produce an *R*_*0*_ = 1.27, still lower than estimated COVID-19 values for *R*_*0*_, but reflective of the mandated restrictions within Australia at those times, and also limiting the very large sizes that can be produced. Using the mutational probability parameters for Delta, a simulated outbreak for this variant produced a clonal network over 24 generations shown in Fig 5. With only these assumptions on the distributions for transmission numbers and mutations, the clonal sizes follow a power law as observed in the actual outbreaks.

**Fig 5:**
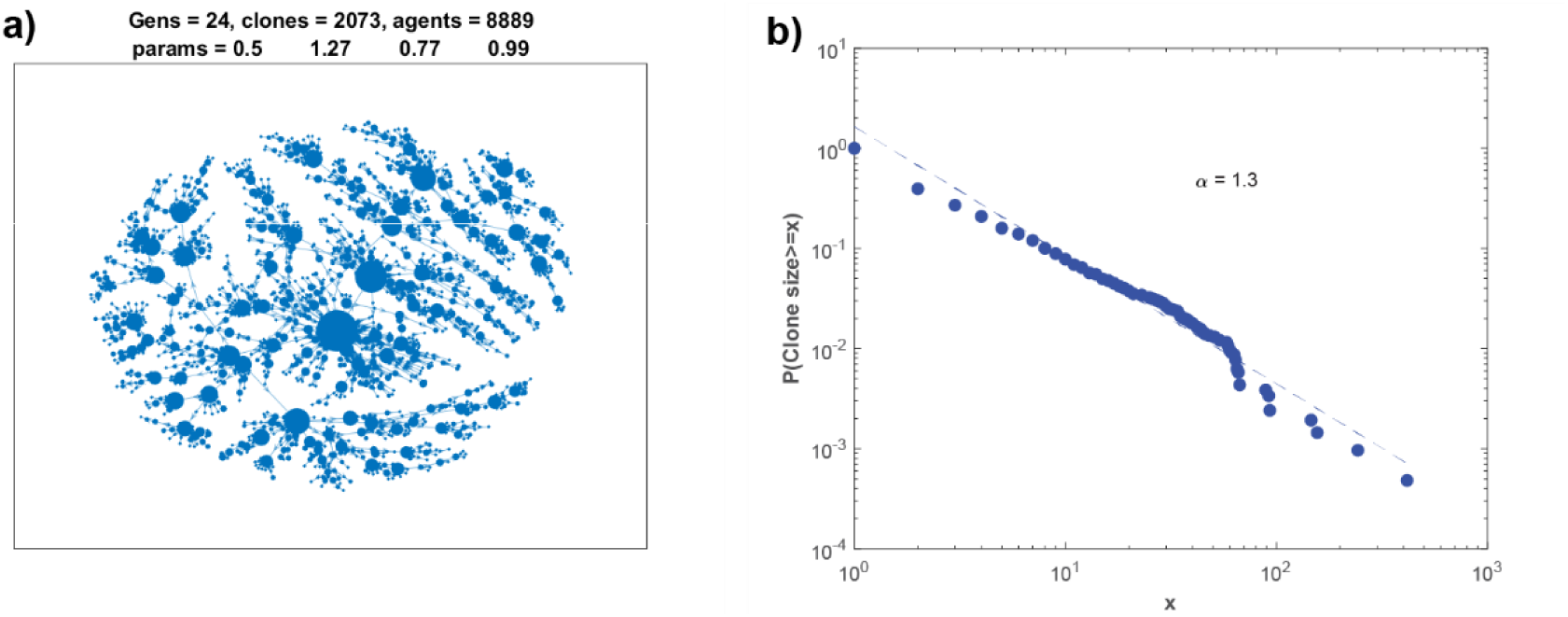
a) simulated Delta clonal transmission network where at each generation (to a maximum of 24 or approximately 8,000 agents) the number of new infections per agent were drawn from a negative binomial distribution with R = 1.27, p = 0.5, mean 1.27, and similarly for the number of mutations per transmission (R = 0.99, p = 0.77, mean 0.30). The size of the nodes represents the square root of the number of sequences within that clone. b) Power law clonal size distribution and fitted regression line α = 1.3.

### Connectedness between groups and recombination

Many of the different groups within a lineage occurred over the same time frame and most likely arose from mutation of a sequence in one group to produce a viral genome containing an insertion/deletion (indel) thereby establishing another group based on ORF lengths.

Multiple alignment of the reference sequence and the root sequences from the 15 different groups in the AY.39.1 lineage uncovered a number of indels including a 6nt deletion within S (start nt posn 22,028) for 11 of the groups, and a 17nt deletion within 7a (start posn 27,608) for a partially overlapping set of 11 groups. The mutations that gave rise to these deletions at exactly the same positions should be independent events such that if the 6nt deletion occurred first then the 17nt deletion should occur in only one of the subsets. However this was not the case. Groups 6 and 9 exhibited a 6nt deletion, while groups 7 and 8 did not; on the other hand, groups 7 and 9 exhibited a 17nt deletion while groups 6 and 8 did not, indicating a recombination event occurred between groups 6 and 7 to produce the combined deletions in group 9. Similar indel patterns suggested recombination also occurred within the Omicron groups. Compared to the reference sequence, Omicron groups 19 and 21 exhibited a 3nt deletion at 6513 (within 1a) but groups 13 and 20 did not; while at position 28370 in N, groups 13 and 19 (and 17 other groups) exhibited an 8 or 9nt deletion while there were no deletions in groups 20 and 21. So group 20 contained no deletions at these locations while groups 13 and 21 contained one of each, while 19 had both deletions.

### Mutational pathways

The transmission networks for each group allow the calculation of how the virus mutates as it is transmitted between individuals. The pathways in each of these groups that contain the most mutations between the very first clone (the root node) to any of the extremities of the network (the leaf nodes) are shown in red in Fig 2 and Fig 3. The mutations that occur in each clone along those paths are shown in Fig 6. Uncertainty in the sequences, and the resulting inability to exactly determine differences between clones, meant that mutations were not completely cumulative along the pathways.

**Fig 6:**
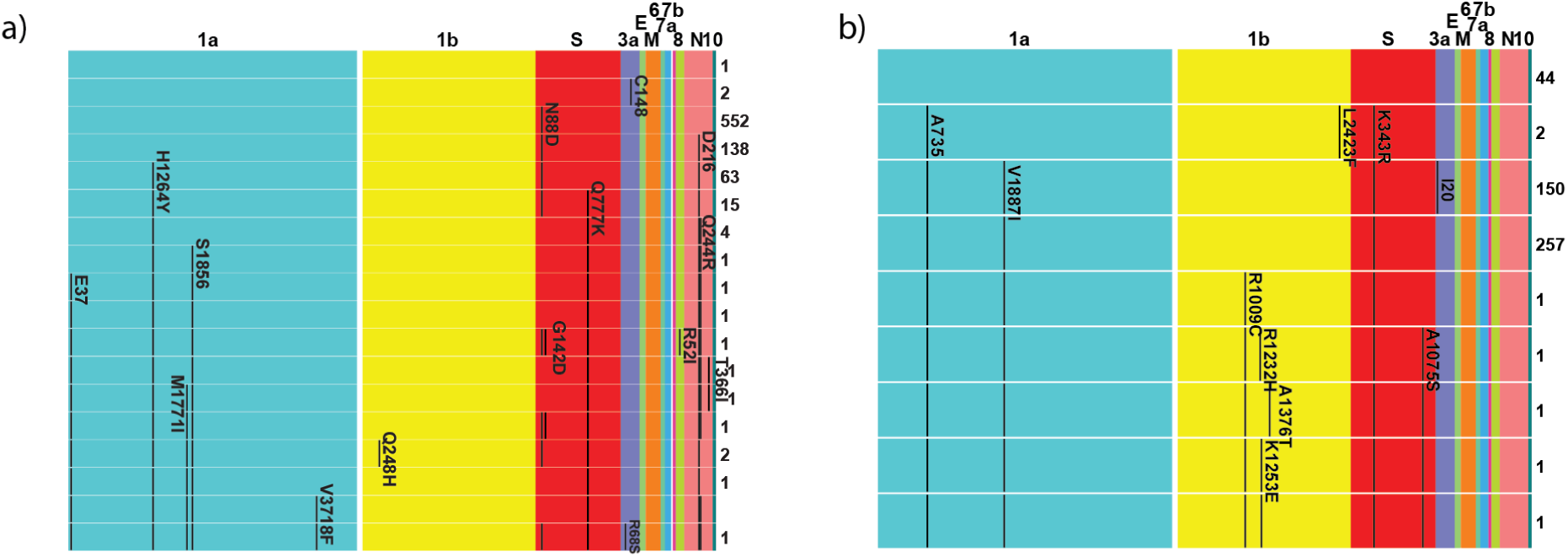
ORF changes along the longest mutational pathways for the a) Delta and b) Omicron groups displayed in Fig 2 and Fig 3. The i ^th^ row represents the i ^th^ clone on the path, with numbers of sequences in that clone recorded on the right. Mutations in the nt sequence are recorded at the amino acid position within the ORF. Synonymous mutations are listed with the residue and position such as D651; nonsynonymous mutations are reported with the changed residue such as K343R. Note that the start of 1b in these calculations was from the start codon ‘ATG’ within that open reading frame, which is 300nt (100AA) after 1b’s commencing sequence of ‘CGGGTTTGCGGTGTAAGT’ (‘RVCGVS’). Numbering in the other ORF can be affected by indels, and these differences are noted in the text. For example, 343 in S for these sequences corresponds to position 346 in the reference sequence.

### Common mutations along pathways

The longest mutational pathways within each lineage/group often shared common mutations. For the Delta groups, either a G142D or D142G appeared in S for 13 of the 31 groups across 6 different lineages. This position is within the spike protein and antibody binding surface (Fig 7a)

**Fig 7:**
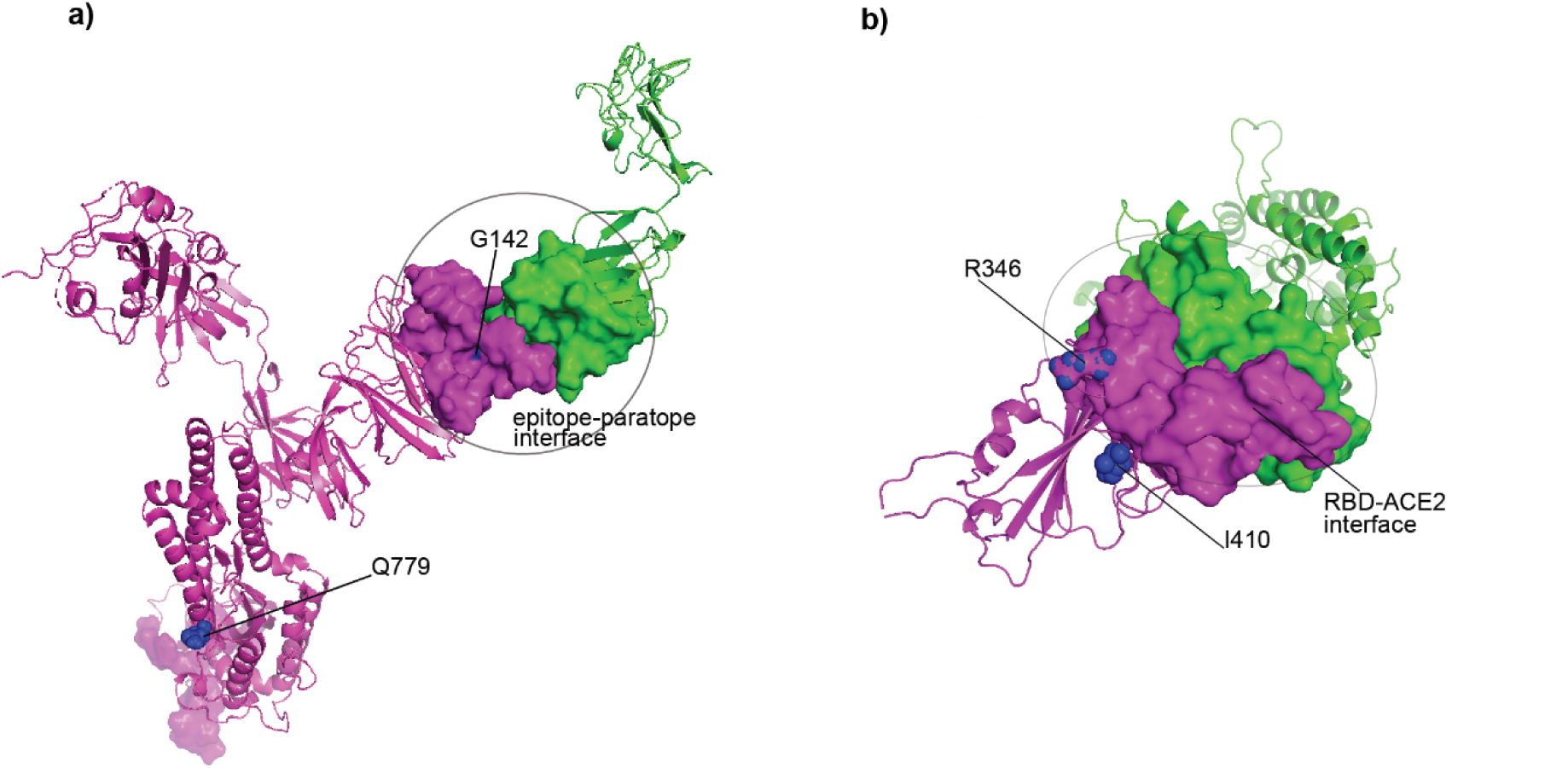
structural locations of the repeated spike mutations for a) Delta (PDB 7LCN), and b) Omicron (PDB 7T9L). The viral spike protein is colored magenta. In a) the antibody heavy chain is colored green; b) ACE2 is colored green The binding surfaces were determined from residues within 8A distance. Within the interface, residues within 5A are in direct physical contact.

The next most frequent mutation was a synonymous mutation in ORFN of D216 for 6 of the AY.39.1 groups. Pairwise Fisher Exact comparisons of the repeated Delta mutations in these pathways only determined significant connections for D216 in ORFN with Q777K (ref. posn 779) in S for two groups (*p=0*.*034*), and D2079E (ref. posn 2179) in 1b with A59V in 3a *(p=0*.*007*). There were no significant connections between the G142D/D142G mutation with any other mutation appearing in these pathways suggesting these occur independently to any other changes.

The Omicron mutations within these pathways were more likely to occur across different groups and show a greater degree of connectedness. An I20 synonymous mutation in 3a was the most frequent, occurring in 17 of the 23 BA.1 groups. The other frequent mutations and their likelihood of appearing in a pathway in the presence/absence of another mutation, are shown in Fig 8. The I20 mutation is less likely to occur in the presence of the A735 and V1887I mutations in 1a, as well as the K343R (ref. posn 346) mutation in the spike protein (Fig 7b), and an L2423F (ref. posn 2523) mutation in 1b. The mutation in the opposite direction for this position, R343K in S, a mutation against which there is reduced neutralizing activity [10], was significantly connected to an I1887V mutation in 1a.

**Fig 8:**
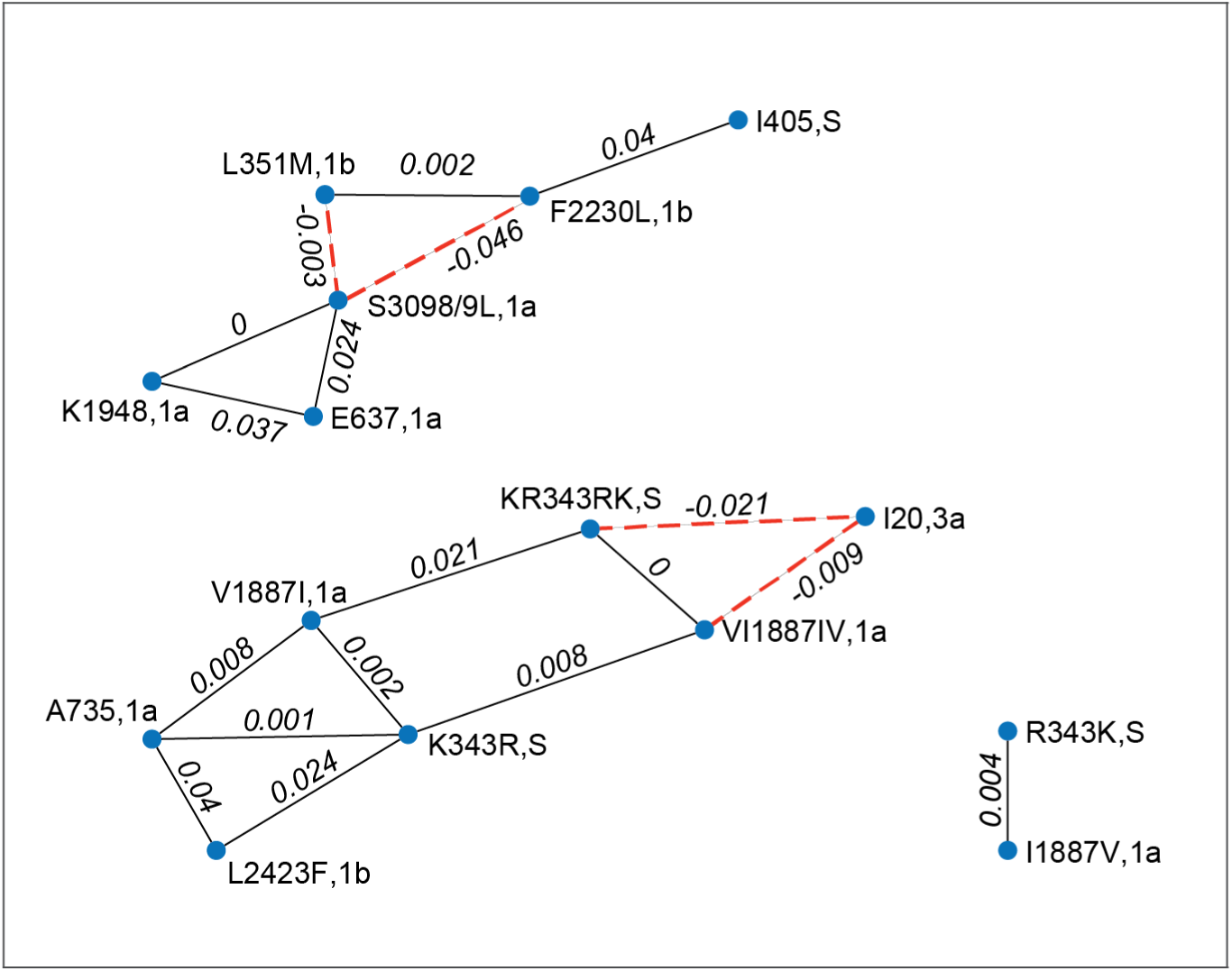
Connectedness between repeated mutations occurring along longest mutational pathways between Omicron groups. Mutations connected by a black edge are more likely to occur together (p values listed on each edge for the Fisher Exact Test), while mutations less likely to occur in the presence of another are shown with red dashed lines connecting them (and the negative of the p value on the edge).

The I20 mutation in 3a was not the only synonymous mutation that was significantly connected to other nonsynonymous ones. The K1948 mutation in 1a appeared in 11 mutational pathways with a S3098L (ref. posn 3099) or S3099L 1a mutation appearing one step further along than K1948 in 10 of these pathways, and in no others, suggesting the synonymous K1948 mutation induced a compensatory mutation S3098/9L. The RNA change within K1948 may have affected subgenomic RNA structures that interact with the viral genome and other host cell RNA, thereby adding selective pressure for the RNA change within S3098/9L [11]. The presence of this last group of mutations (along with E637 in 1a), tended to occur in the absence of another group of mutations L351M (ref. posn 451) and F2230L (ref. posn 2330) in 1b and I405 (ref. posn 410) in S, suggesting a different escape pathway for these groups. It is of interest that these mutational groupings include changes in multiple ORF.

## Discussion

Using SARS-CoV-2 sequencing data from both Delta and Omicron variants, probable transmission networks have been estimated. The analysis not only shows the high-level expansion of many clones within an outbreak, but also the degree to which the virus mutates even in these relatively small outbreaks by global standards. Each node in the clonal network represents a new mutation and the edges in each pathway leading from the root node show successive mutations, which if fitter against immune and vaccine coverage, lead to greater likelihood of generating a variant of concern.

Our analysis estimated a mean number of 1 and 0.79 nt mutations per transmission for Delta and Omicron respectively. The few virions that establish infection and the low probability of variant transmission during acute infection [2, 12, 13], especially for Omicron with its shorter serial interval [14], still give rise to large clone sizes as observed here. Many of those variants will die out, because those individuals have low contact rates, or host gene variants may reduce cellular entry and viral replication [15], or the variant is less fit. Expansion of some clones is likely due to a combination of super-spreader events and variant fitness [16], an investigation of which would require further epidemiological and genetic analysis.

The mutational pathways observed in each group can indicate which of the mutations offer advantages in their spread. Although the residue repeatedly switched between a G and D at position 142 in the spike protein along these pathways, it seemed to offer no selective advantage for Delta. The G142D mutation has been associated with increased viral load and immune evasion, exhibiting frequent back mutation as observed here [17]. This same mutation was also associated with symptomatic relapse in a kidney transplant patient who was unable to clear the virus [18]. It may be that the presence of this mutation is associated with immune pressure and possibly escape. However, the frequent back mutations indicate that there is a loss of fitness in some other aspect of the viral life cycle. The Omicron S protein mutations of positively charged R and K basic residues at position 343 (position 346 in the reference sequence), have epitope and attachment functions, playing a role in ACE2 binding and immune escape [19]. The Omicron mutational pathways uncovered more consistent changes that were linked to other mutations some of which occurred in different ORF (Fig 8). Some of these appearances may be due to different infections being established in Australia when border controls were relaxed in late 2021, and the cases being incorrectly amalgamated in the one network. This may have been the case for the A735,1a, V1887I,1a, L2423F,1b, K343R,S, set of mutations[10] that mostly (but not always) appeared in the same mutational step. However, this is not likely to be the only reason. The 3a synonymous mutation I20 for instance occurred at steps in the pathway that were consistent with the appearance of other mutational steps both before and afterwards, which would be unlikely if the I20 mutation originated from artificially linking sequences containing this mutation with those that did not. This synonymous mutation was not the only one that appeared consistently and indicated some benefit to the virus in its transmission. The K1948 mutation consistently preceded an S3098/9L mutation, both in ORF1a. Although synonymous mutations leave the associated protein unaffected, they can influence the structure of the many subgenomic RNA used in the infection cycle of SARS-COV-2 [11]. Whatever advantage these mutations provide, they do so through non-spike alterations.

SARS-COV-2 arose from recombination [20], so it is reasonable to expect recombination occurs within the epidemic. This has been documented by others including Jackson and colleagues who determined recombinants from mosaic patterns circulating in the UK [21], while the recombination rate has been estimated in excess of 4 × 10^−5^ per nt site per year [22]. The much smaller outbreaks within Australia also showed evidence of recombination in both the Delta and Omicron variants. These events were determined from patterns of indels, the creation of which are much rarer than individual mutations (given the extent of some of the transmission networks that retain the same ORF lengths). For that reason, and given the highly conserved nature of the genome within a transmission network, there are likely to be other recombinations that are not detected, many of which would recombine with very similar sequences.

There are many limitations in these calculations, in particular the uncertainties of many nt in the sequences. The Omicron sequences contained more uncertain nt than Delta which made estimation of the ORF domains particularly difficult for many of the groups, resulting in approximations in distances between sequences (and clones), and the inability to assess some of the mutational pathways over all ORF (S2 Fig). The Omicron sequences were only collected in some states for individuals whose treatment was dependent on knowledge of the variant, so the networks and resulting mutation rate estimates would have been impacted.

However, to the best of our knowledge, this is the first method to provide a population level transmission network for any virus based solely on data, something that has only been possible due to the huge investment in viral sequencing throughout the COVID-19 pandemic. Contact tracing efforts have limited scalability in large outbreaks, while viral sequencing efforts are becoming more popular and more rapid. By using the method developed here, it may be possible to complement traditional contact tracing efforts for not only this virus, but for any infectious diseases where sequencing can be used to link cases.

In conclusion, we provide a data driven model of SARS-CoV-2 transmission networks. We observed relatively high mutation and recombination, highlighting the need for ongoing vigilance and research into future escape variants. Further, the model itself may have practical applications in future outbreaks of SARS-CoV-2. This work can be readily applied to other viruses where large scale sequencing is available.

## Methods

Complete sequences for Oceania/Australia were downloaded from GISAID for the variant VOC Delta GK, submission to 31 October 2021, and variant VOC Omicron GRA, submission to 11 January 2022. After separation into their lineages, sequences were pairwise aligned to the reference sequence EPI_ISL_412026 [9], after which the start and end of the 12 ORFs were estimated. Rather than determining the start of 1b at position 13468 in the reference sequence and beginning with ‘CGGGTTTGCGGTGTAAGT’ and AA sequence ‘RVCGVS’, the open reading frame commencing with the start codon ‘ATG’ was determined and this with its corresponding stop codon was used for ORF1b. In all cases this occurred 300nt (100AA) later. Sequences were separated into groups based on unique ORF lengths. Estimation of the ORF domains was necessary for tracking synonymous/nonsynonymous mutations within each ORF. Due to the high level of uncertain nt for Omicron sequences, their grouping was restricted to lengths of ORF 1a, 1b, 3a, 6, 7a, 10, and distances between the end of 1b and the start of 3a, the end of 3a and the start of 6, and the end of 10 and the start of 1a. Groups containing fewer than 10 sequences were discarded.

After sorting sequences by increasing number of uncertain nt, they were separated into clones based on zero pairwise distance and the same distance between all sequences in separate clones. This latter aspect meant some sequences in separate clones had zero mutations between them (but they were different distances to sequences in at least one other clone). A directed graph *G*, was constructed where each clone was represented by a node (size equal to the number of sequences in the clone). Within each clone the sequence with the earliest date of sampling was termed the clone root node. A directed edge from clone *i* to clone *j* was included if the clones were separated by at most 6nt mutations, and for each sequence *k* in clone *i* if its date of sampling *t*_*k*_ was such that the date *t*_*j*_ of the clone *j* root node satisfied *– 5 ≤ t*_*j*_ *– t*_*k*_ *≤ 17*. Limiting incoming edges to the clone root node ensured that the transmission to a new clone occurred at the first time that clone was sampled. The probability of a transmission occurring between two nodes was based on a Gamma distribution *p(t*_*j*_ *– t*_*k) =*_ *Γ(t*_*j*_ *– t*_*k*_ *– 10*.*5)* with shape and scale parameters of 21.96 and 1.57 [6]. With these values *p(–5) ≈ p(17)*. For transmission between clones separated by *d* mutations, the edge weight was modified by a penalty term, *w= – log p(t*_*j*_ *– t*_*k*_*) + Md*. The maximum value at *t* = 17 was *– log p ≈ 9*. Setting *M* = 10 for a single mutation resulted in a weight exceeding any sampling time difference; calculations with *M* = 2, equivalent to an additional 9 day time difference, were similar but tended to produce mutational pathways that were less consistent in maintaining a mutation once it arose. The maximum of 6nt mutations for an edge was chosen since most sequences were clonal suggesting a low mutation rate, and also to limit the combinatorial growth in edge numbers with higher mutational distances. Even with this limit, the starting graph can be very large. For example the within-clone graph for Fig 4a began with 630 nodes and 286,980 edges.

Weakly connected components for the graph of each group were extracted. For each connected component of size at least 10, the minimal arborescence was calculated using the method of Chu, Liu and Edmonds [7, 8].

All computations were performed with Matlab R2021b (The Mathworks Inc., Natick MA, USA).

## Data Availability

All sequence data are available from GISAID.

https://www.gisaid.org

## Acknowledgements

This work was funded by an Australian Research Council Discovery grant (DP180103893), awarded to J.M.M. The Australian Research Council (arc.gov.au) had no role in study design, data collection and analysis, decision to publish, or preparation of the manuscript. We gratefully acknowledge in Supplementary Table S3, the authors from the Originating Laboratories responsible for obtaining the specimens and the Submitting Laboratories where genetic sequence data were generated and shared via the GISAID Initiative, on which this research is based.

## Supporting information captions

S1 Fig. The optimal transmission networks for all Delta groups showing the path with the most mutations as well as their occurrence in the 12 ORF.

S2 Fig. The optimal transmission networks for all Omicron groups showing the path with the most mutations as well as their occurrence in the 12 ORF.

S3 Fig. Power law distributions of clone size.

S4 Fig. Number of mutations per transmission for the largest Delta clonal network.

S5 Fig. Number of mutations per transmission for the largest Omicron clonal network.

S1 Table: Probability of mutation per transmission, power exponent of clone size, and distribution of number of mutations per transmission for Delta.

S2 Table: Probability of mutation per transmission, power exponent of clone size, and distribution of number of mutations per transmission for Omicron.

S3 Table: GISAID Acknowledgement

## Author contributions

J.M.M and E.H.A performed the sequence analysis. J.M.M coded the network analysis. All authors contributed to the article preparation.

## Data availability

All sequence data are available from GISAID and accessed as described in Methods.

## Code availability

Code to create transmission networks from sequence data, and to produce the agent-based model, are available at https://github.com/john1murray1m/SARS-COV-2-Transmission-Networks.

