## Supplementary Figures and Tables for "SARS-COV-2 Delta and Omicron community transmission networks"

### S1 Fig

The optimal transmission networks for all Delta groups showing the path with the most mutations as well as their occurrence in the 12 ORF

AY.39.1, grp = 2  1  1, no. clones = 23, no. seqs = 38, 02-Sep-2021

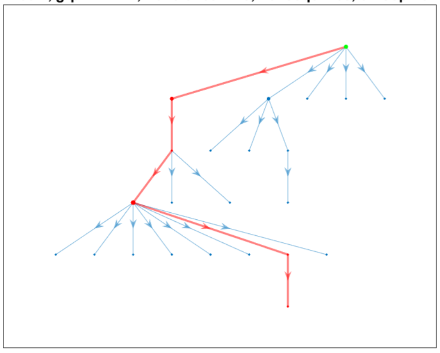

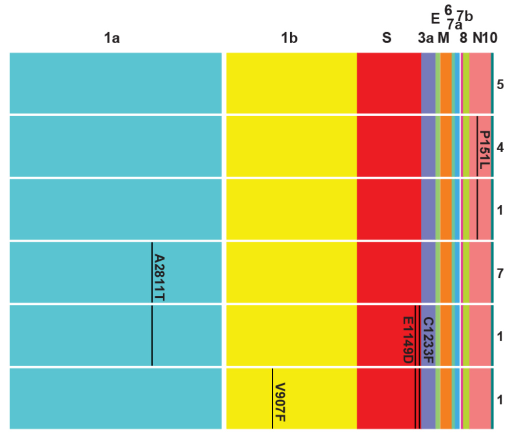


AY.39.1, grp = 2  2  1, no. clones = 19, no. seqs = 26, 07-Sep-2021

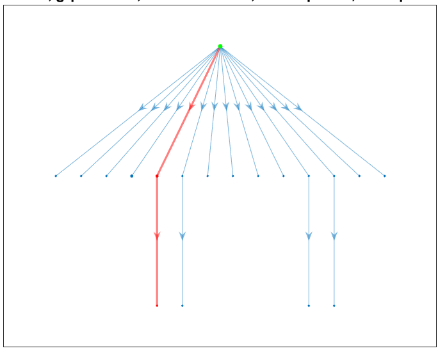

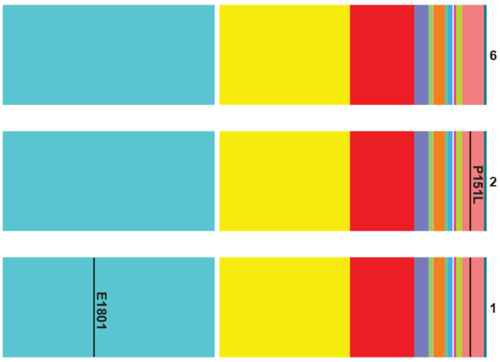


AY.39.1, grp = 2  3  1, no. clones = 18, no. seqs = 26, 01-Sep-2021

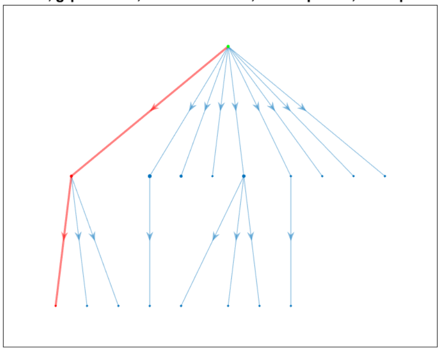

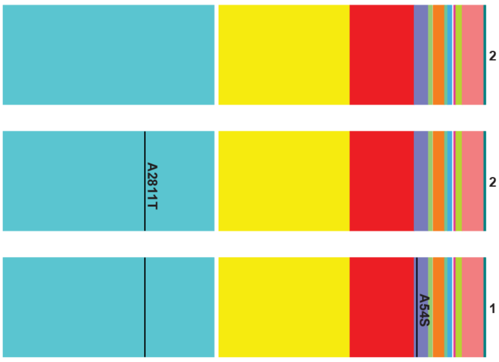


AY.39.1, grp = 2  4  1, no. clones = 487, no. seqs = 908, 23-Jun-2021

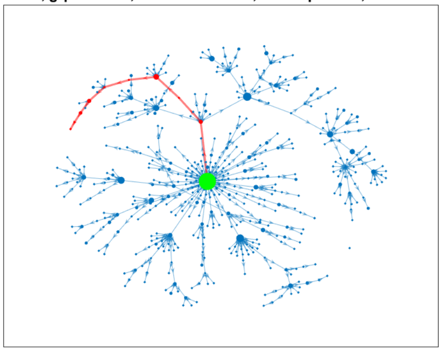

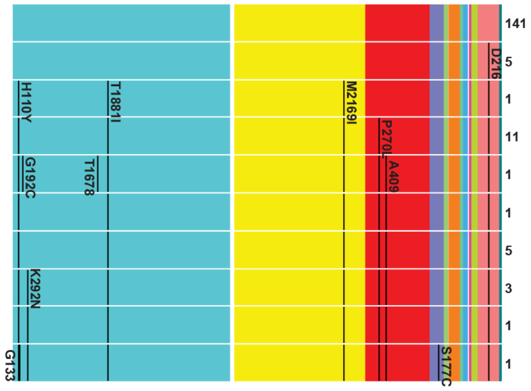

AY.39.1, grp = 2 5 1, no. clones = 22, no. seqs = 37, 02-Sep-2021

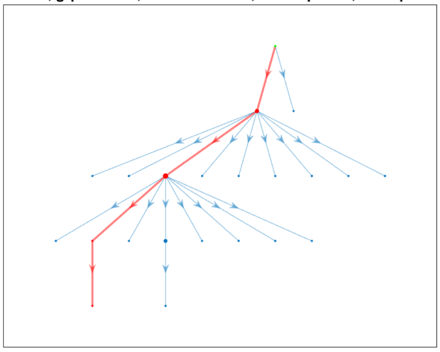

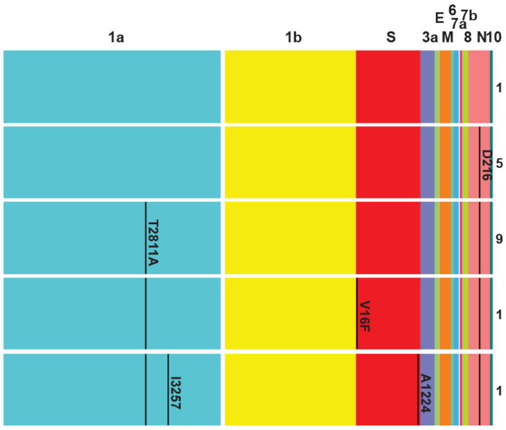


AY.39.1, grp = 2 6 1, no. clones = 107, no. seqs = 198, 24-Jun-2021

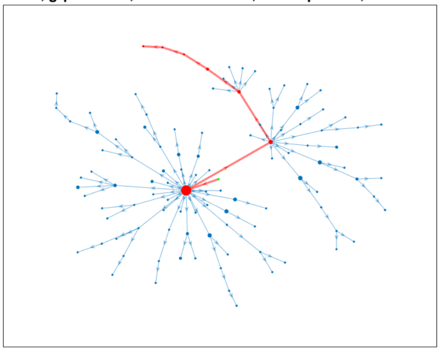

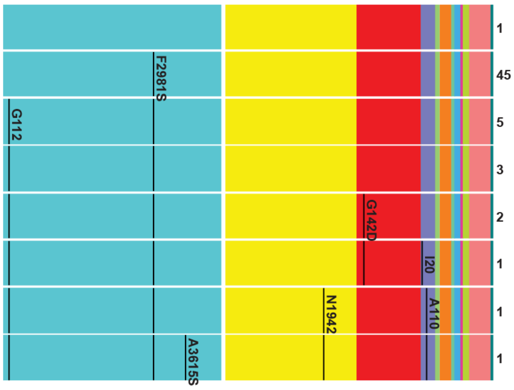


AY.39.1, grp = 2 7 1, no. clones = 161, no. seqs = 264, 30-Jun-2021

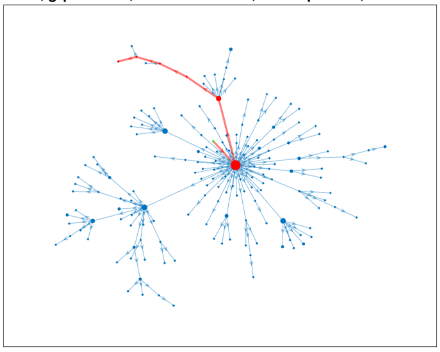

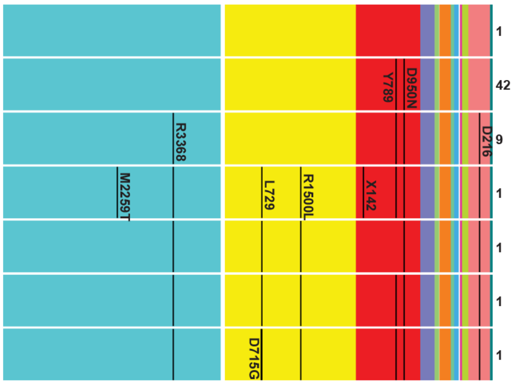


AY.39.1, grp = 2 8 1, no. clones = 51, no. seqs = 64, 04-Jul-2021

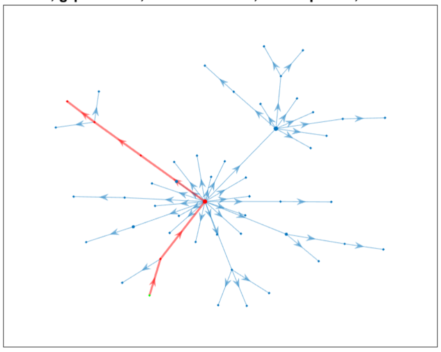

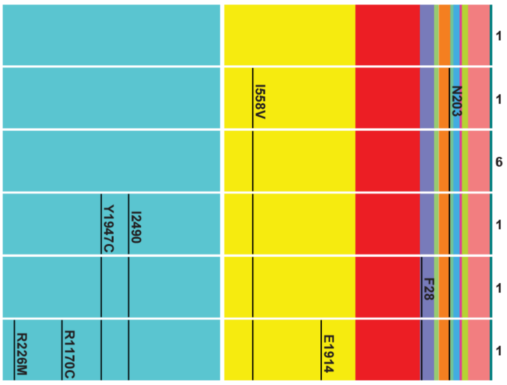

AY.39.1, grp = 2  9  1, no. clones = 17, no. seqs = 21, 03-Aug-2021

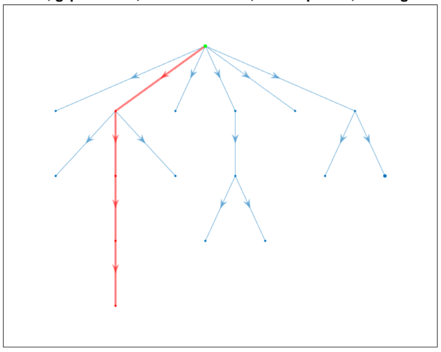

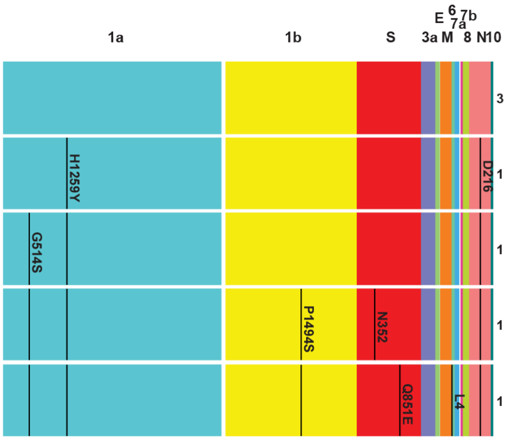


AY.39.1, grp = 2  10  1, no. clones = 14, no. seqs = 22, 11-Jul-2021

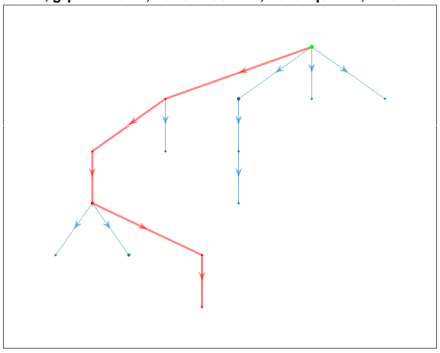

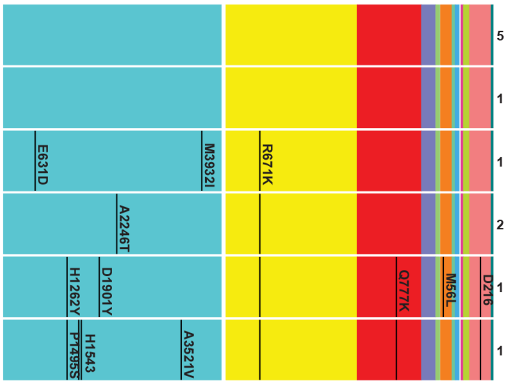


AY.39.1, grp = 2  11  1, no. clones = 2176, no. seqs = 6136, 13-Jun-2021

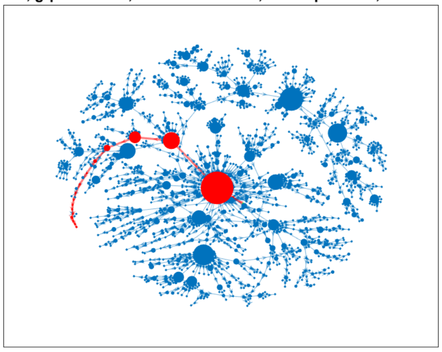

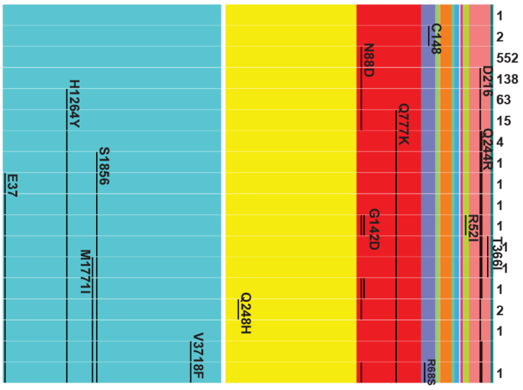


AY.39.1, grp = 2  12  1, no. clones = 241, no. seqs = 721, 17-Aug-2021

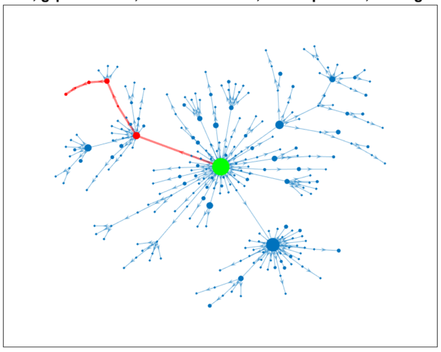

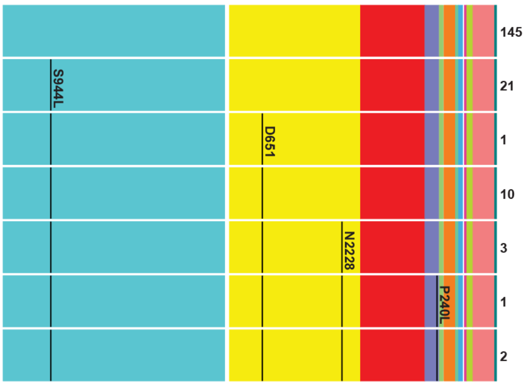

AY.39.1, grp = 2  13   1, no. clones = 479, no. seqs = 1282, 16-Jun-2021

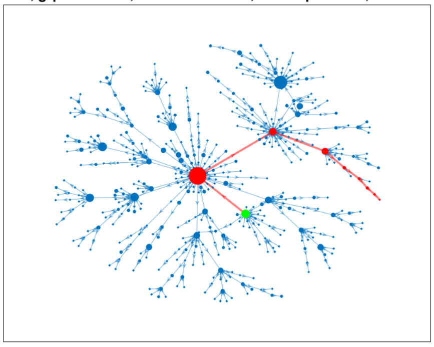

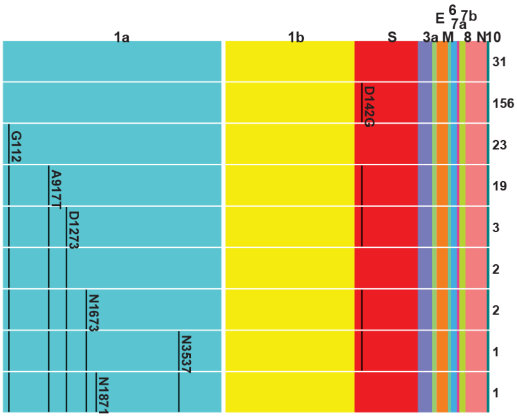


AY.39.1, grp = 2  14   1, no. clones = 238, no. seqs = 591, 24-Jun-2021

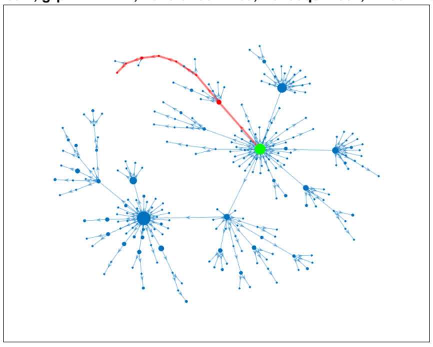

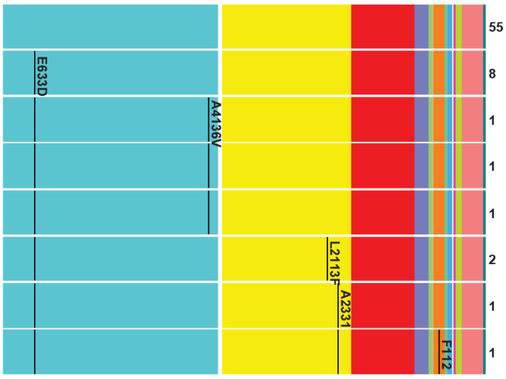


AY.39.1, grp = 2  15   1, no. clones = 50, no. seqs = 103, 20-Jun-2021

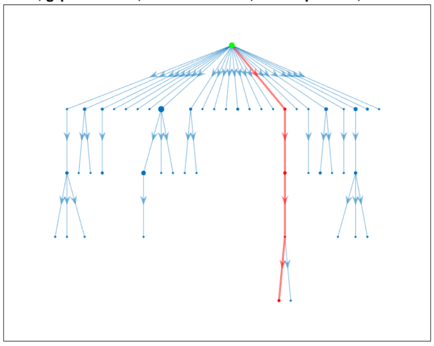

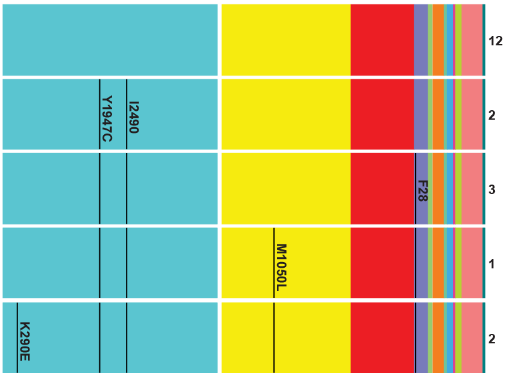

B.1.617.2, grp = 1  1  1, no. clones = 21, no. seqs = 33, 03-Apr-2021

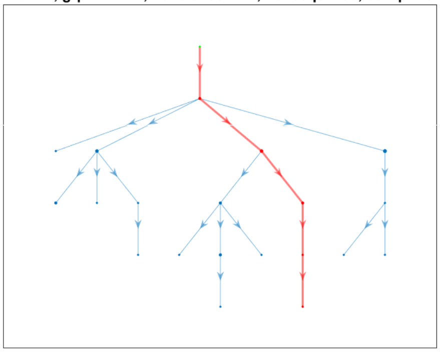

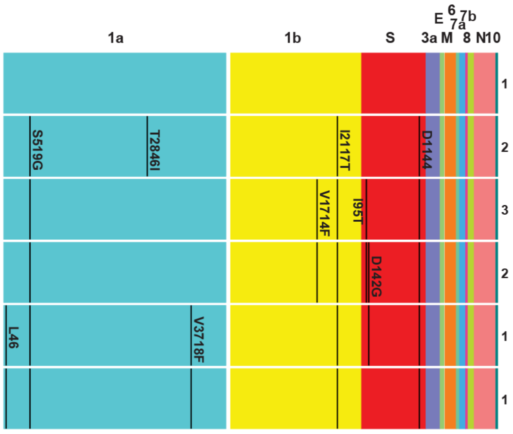


B.1.617.2, grp = 1  1  2, no. clones = 12, no. seqs = 18, 13-Apr-2021

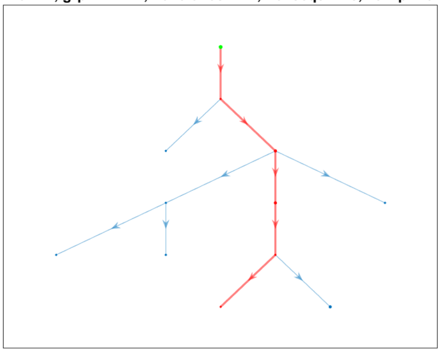

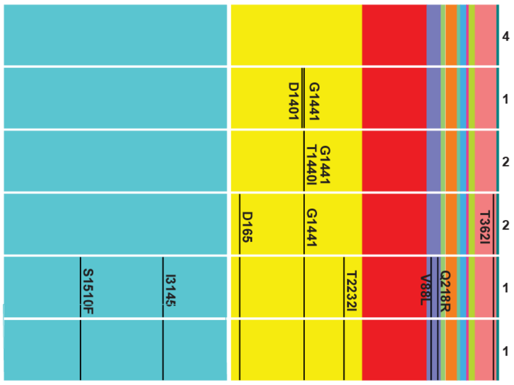


AY.23, grp = 4  1  1, no. clones = 19, no. seqs = 39, 11-Jun-2021

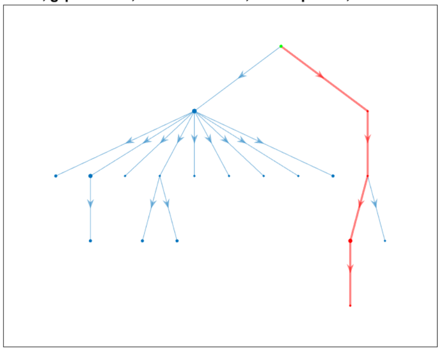

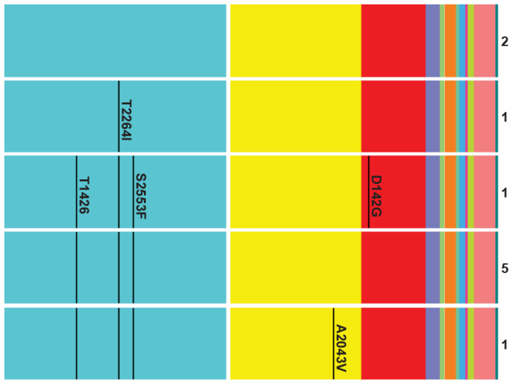

AY.39, grp = 5  1  1, no. clones = 27, no. seqs = 35, 27-Aug-2021

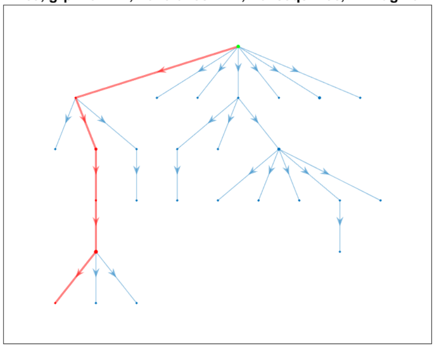

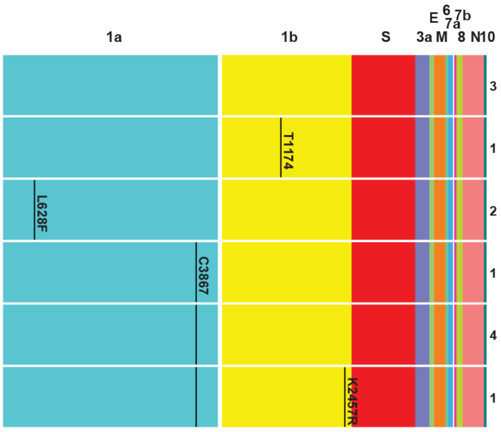


AY.39, grp = 5  2  1, no. clones = 166, no. seqs = 371, 09-Jul-2021

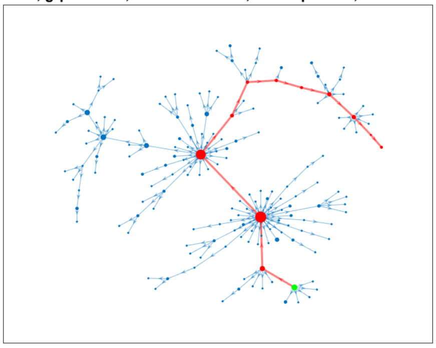

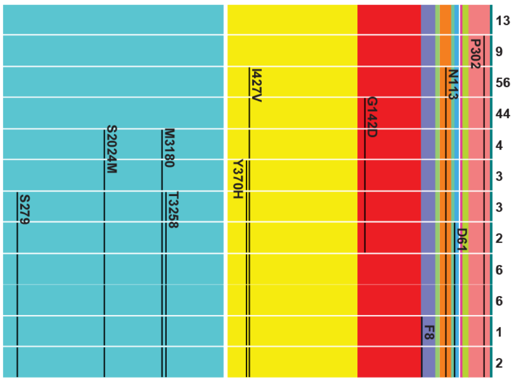

AY.39.1.3, grp = 6 1 1, no. clones = 110, no. seqs = 155, 28-Jul-2021

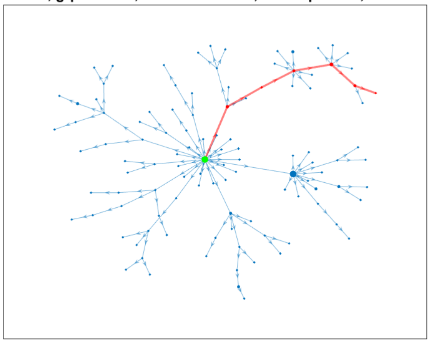

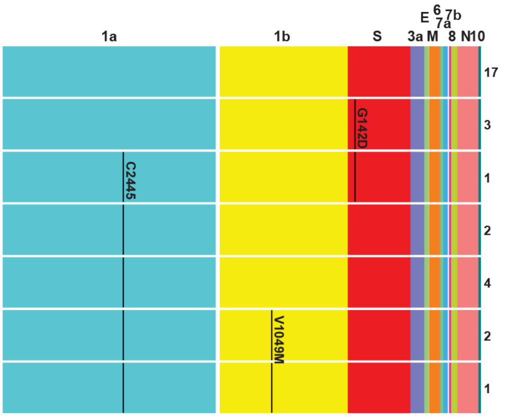


AY.39.1.3, grp = 6 2 1, no. clones = 45, no. seqs = 62, 23-Jul-2021

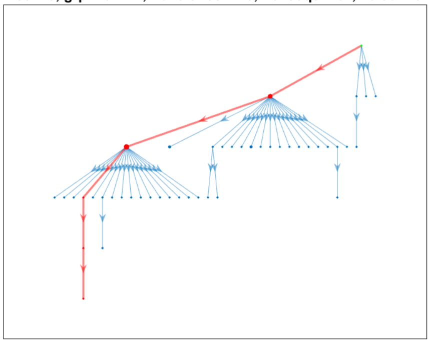

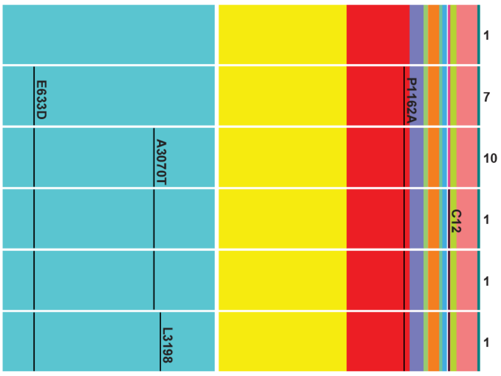


AY.39.1.3, grp = 6 3 1, no. clones = 463, no. seqs = 1311, 25-Jul-2021

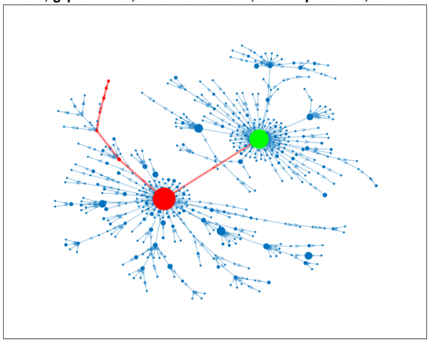

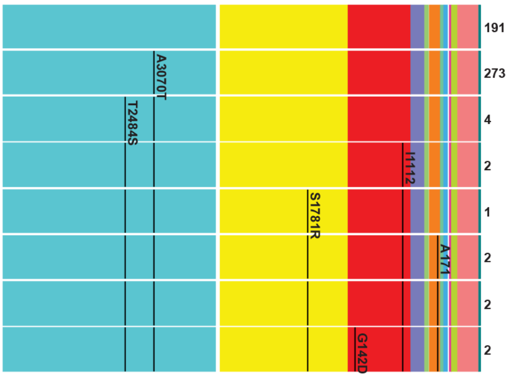


AY.39.1.3, grp = 6 4 1, no. clones = 49, no. seqs = 103, 29-Jul-2021

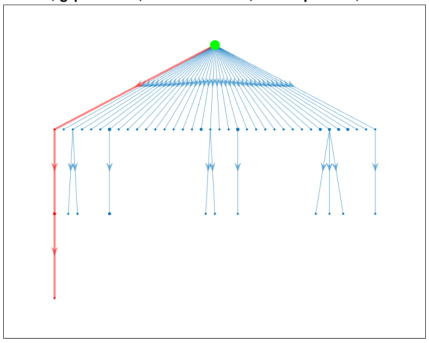

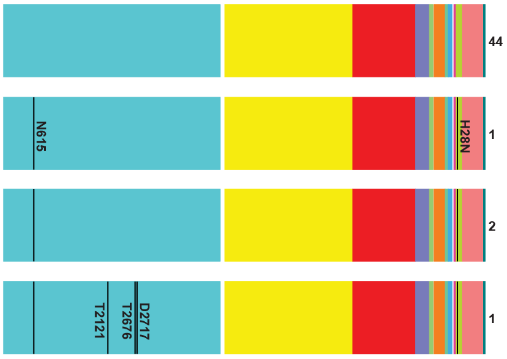

AY.39.1.2, grp = 7 1 1, no. clones = 299, no. seqs = 392, 20-Jul-2021

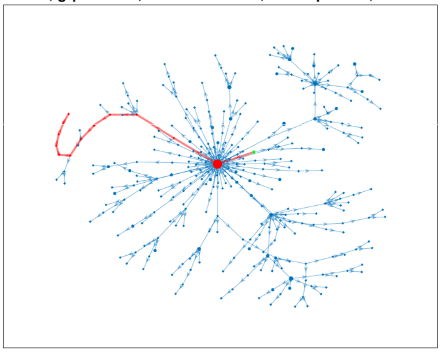

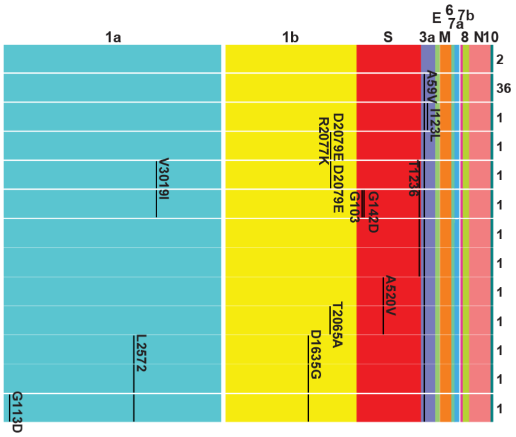


AY.39.1.2, grp = 7 2 1, no. clones = 93, no. seqs = 115, 09-Aug-2021

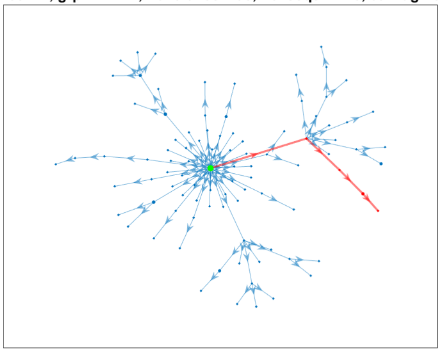

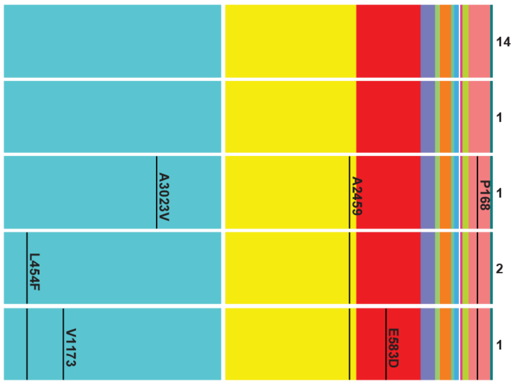


AY.39.1.2, grp = 7 3 1, no. clones = 19, no. seqs = 24, 31-Aug-2021

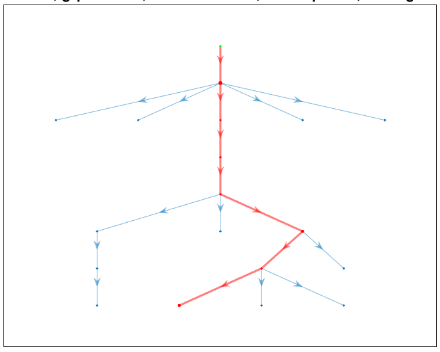

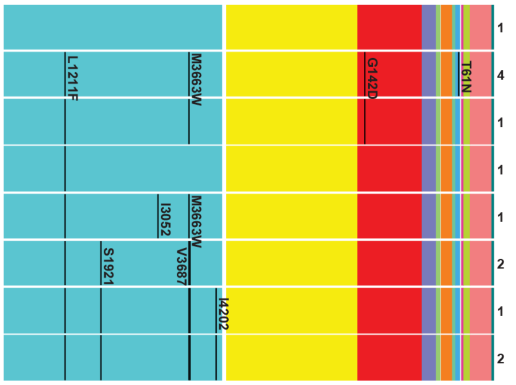


AY.39.1.2, grp = 7 4 1, no. clones = 13, no. seqs = 33, 25-Aug-2021

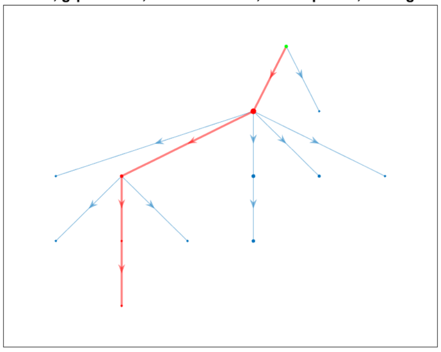

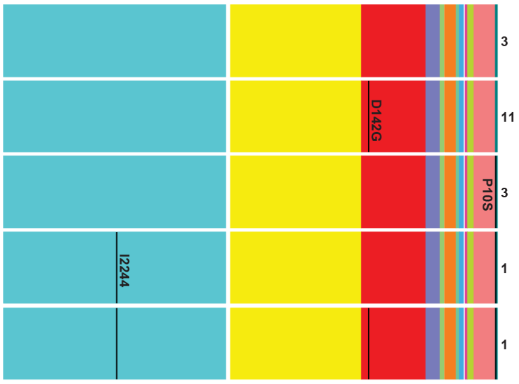

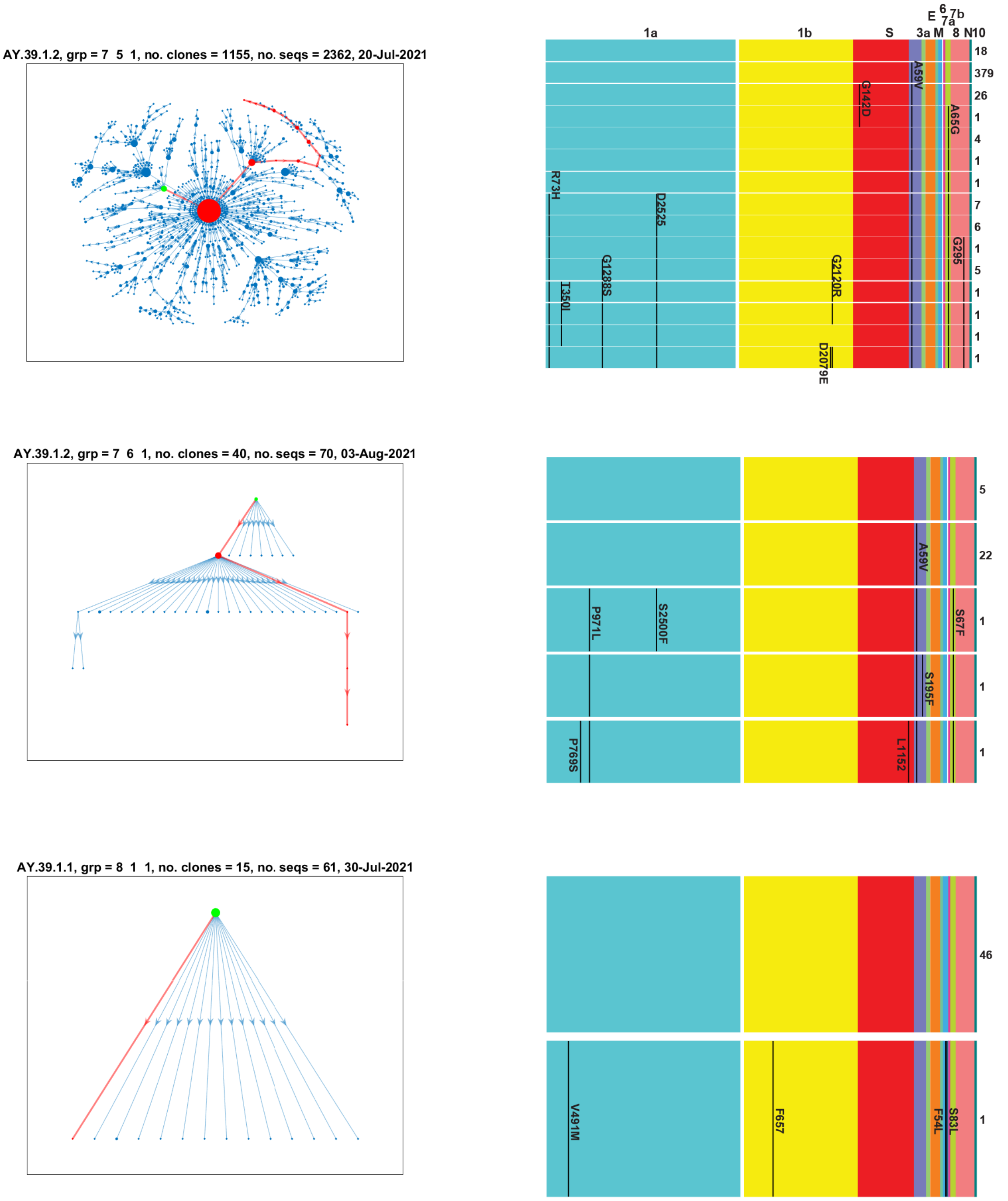


S1 Fig: Estimated clonal transmission networks for all Delta groups, the longest mutational pathways (red), and the resulting ORF mutations measured between each clone on the path and the root node (green). The notations above the networks show the lineage, the group numbers within the lineage, the number of clones and sequences included in the network, and the date associated with the root node. The mutational steps for the longest mutational pathway are noted as AA and positions within each ORF (measured from the start of the ORF in that group rather than in terms of the reference sequence), and where these appear in each step as shown by the extent of the black lines. Numbers to the right at each step denote the number of sequences within each clone. A mutation not present in a subsequent step may be due to its loss, or the root node in that clone contains 'N's at those positions.

### S2 Fig

The optimal transmission networks for all Omicron groups showing the path with the most mutations as well as their occurrence in the 12 ORF

BA.1, grp = 1  1, no. clones = 12, no. seqs = 13, 29-Dec-2021

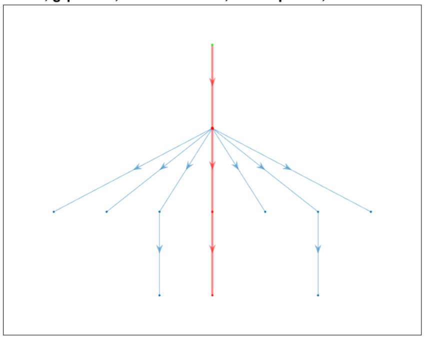

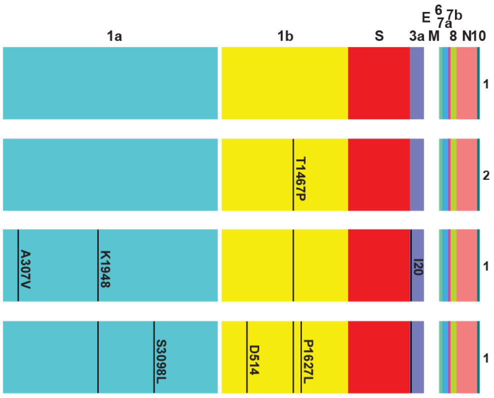


BA.1, grp = 2  2, no. clones = 24, no. seqs = 28, 06-Dec-2021

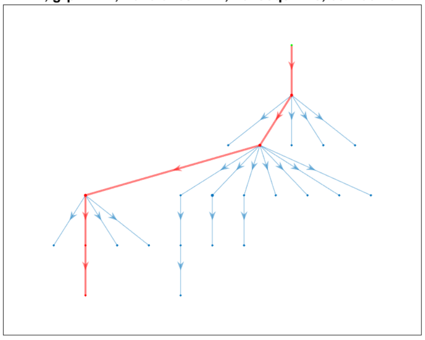

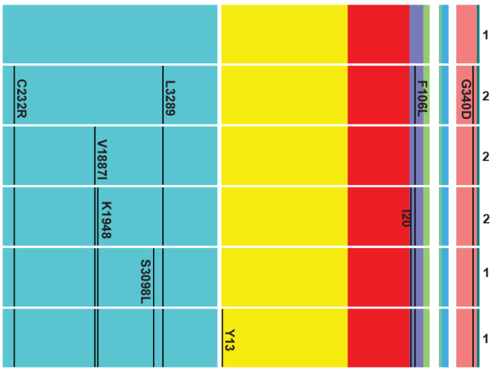


BA.1, grp = 3  3, no. clones = 37, no. seqs = 60, 20-Dec-2021

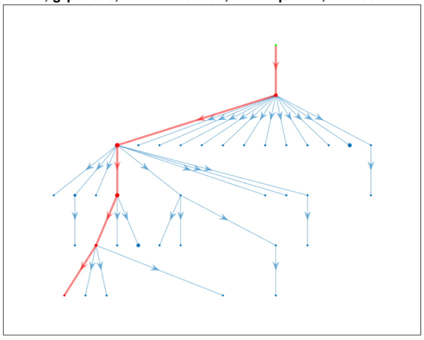

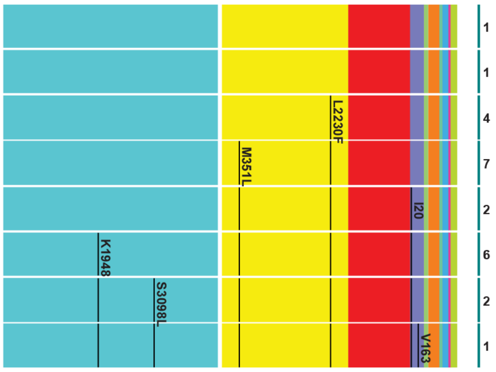


BA.1, grp = 4  4, no. clones = 25, no. seqs = 50, 19-Dec-2021

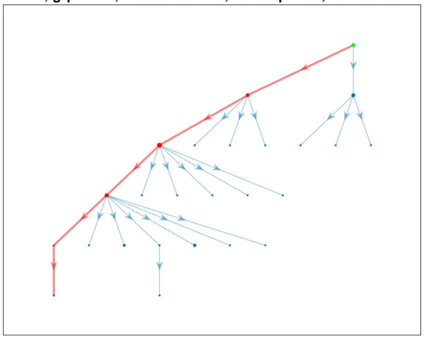

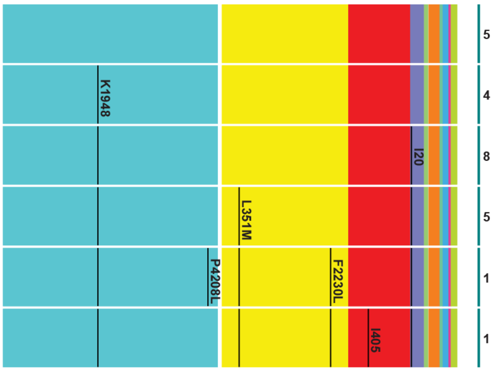

BA.1, grp = 4  4, no. clones = 10, no. seqs = 10, 24-Dec-2021

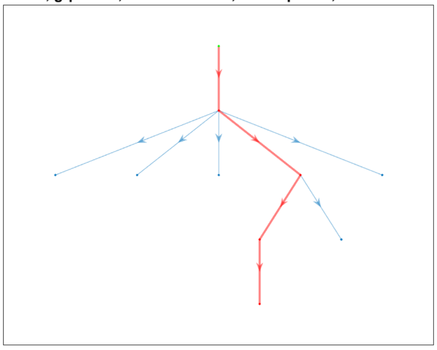

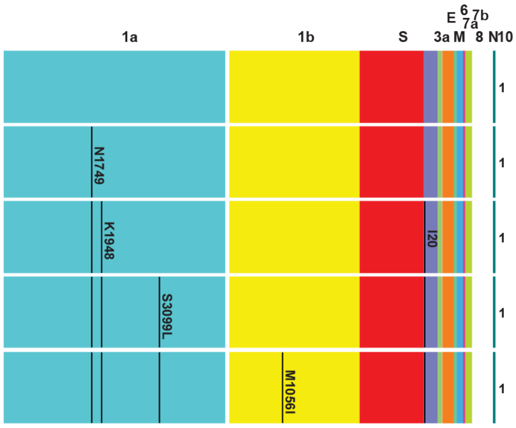


BA.1, grp = 5  5, no. clones = 21, no. seqs = 49, 19-Dec-2021

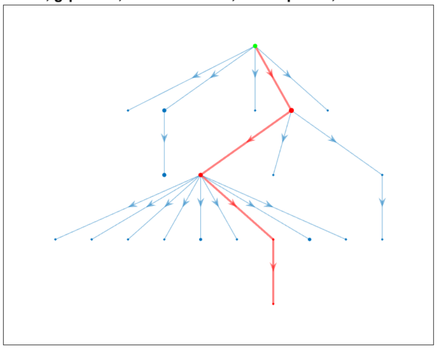

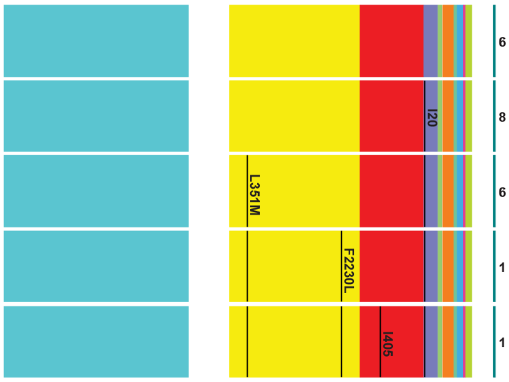


BA.1, grp = 6  6, no. clones = 542, no. seqs = 2119, 27-Nov-2021

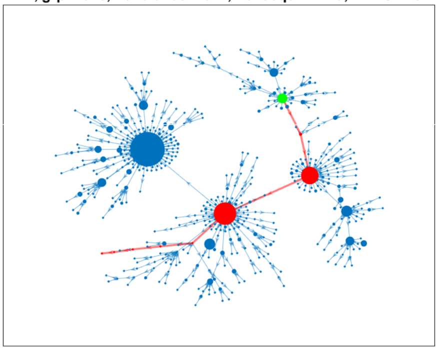

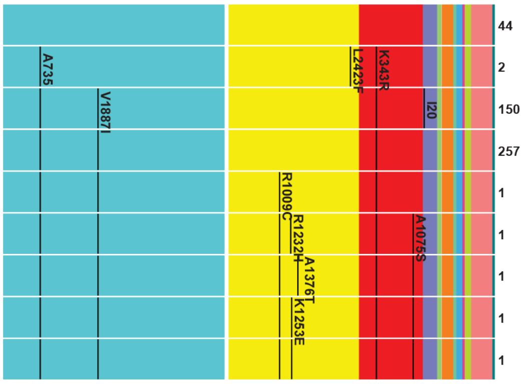


BA.1, grp = 7  7, no. clones = 40, no. seqs = 105, 18-Dec-2021

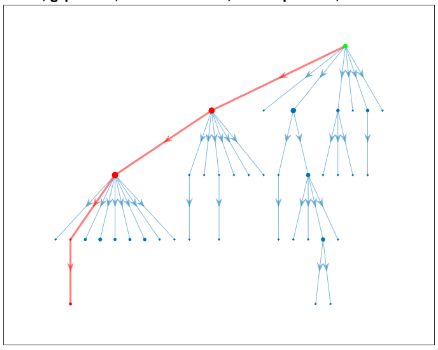

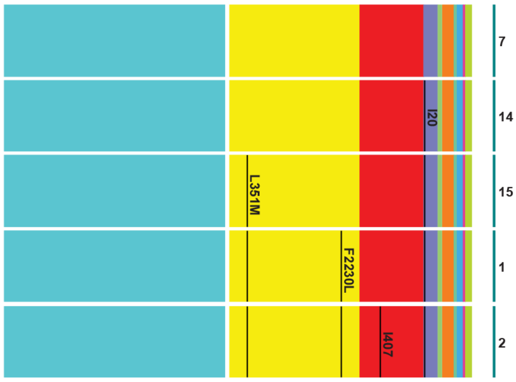

BA.1, grp = 8  8, no. clones = 12, no. seqs = 19, 20-Dec-2021

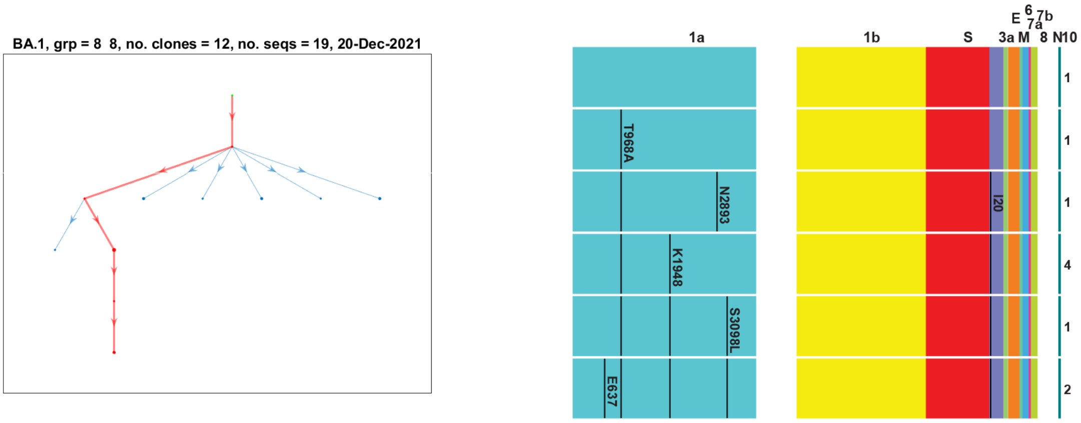


BA.1, grp = 9  9, no. clones = 103, no. seqs = 411, 16-Dec-2021

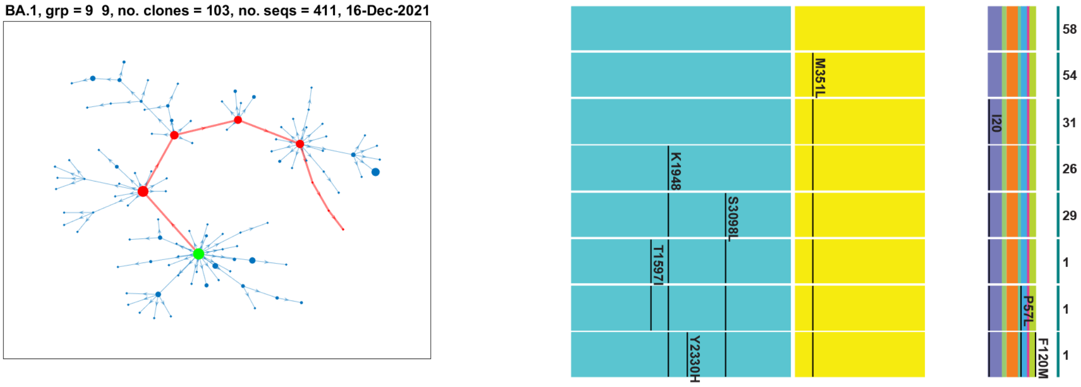


BA.1, grp = 10  10, no. clones = 13, no. seqs = 25, 18-Dec-2021

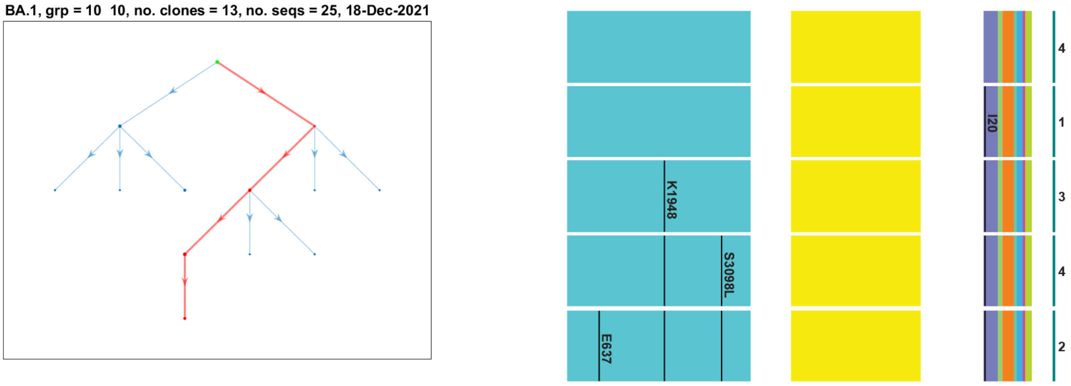


BA.1, grp = 11  11, no. clones = 11, no. seqs = 15, 20-Dec-2021

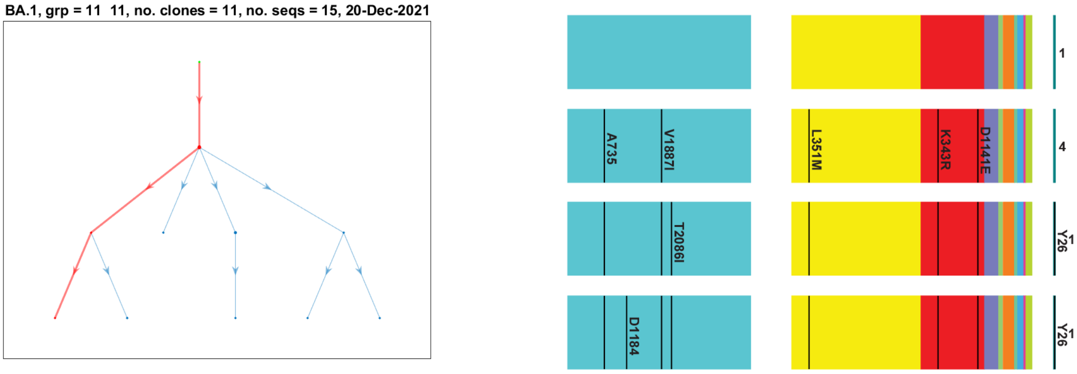

BA.1, grp = 13 12, no. clones = 88, no. seqs = 156, 12-Dec-2021

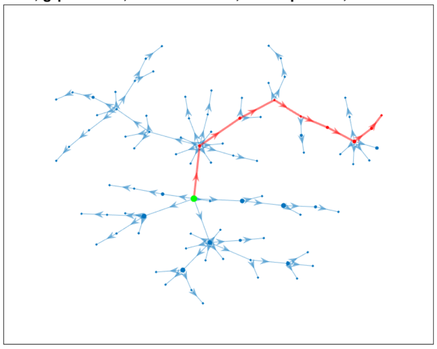

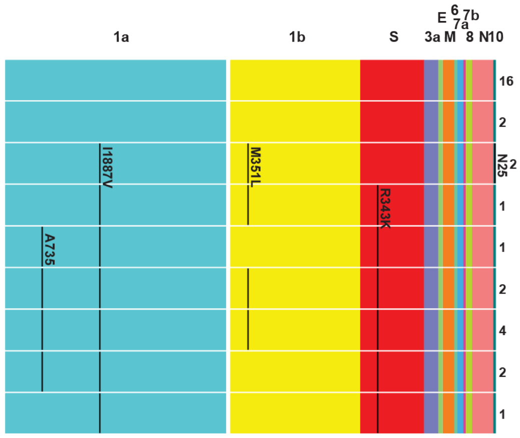


BA.1, grp = 14 13, no. clones = 12, no. seqs = 16, 16-Dec-2021

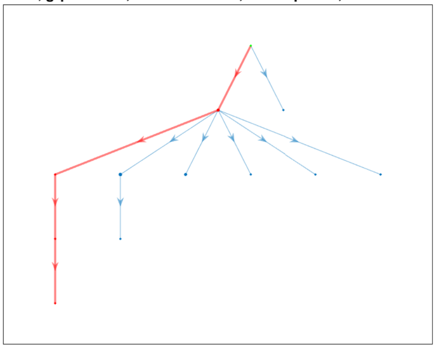

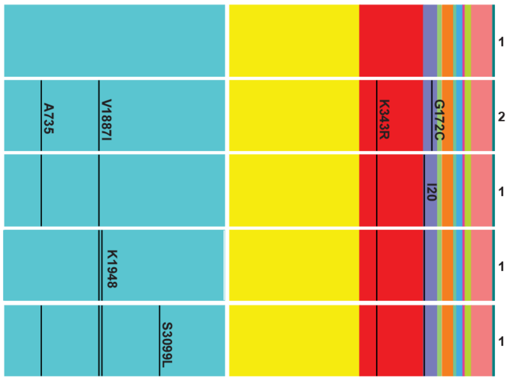


BA.1, grp = 15 14, no. clones = 14, no. seqs = 23, 19-Dec-2021

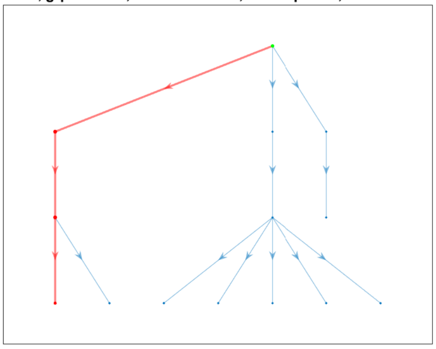

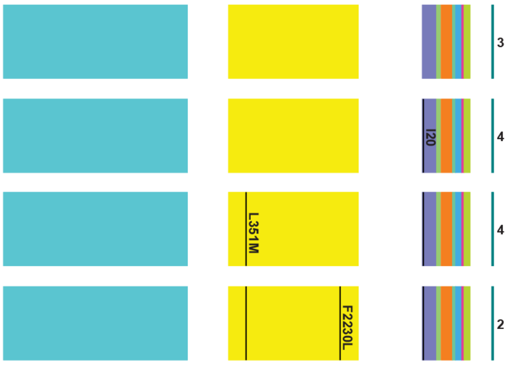


BA.1, grp = 16 15, no. clones = 20, no. seqs = 39, 19-Dec-2021

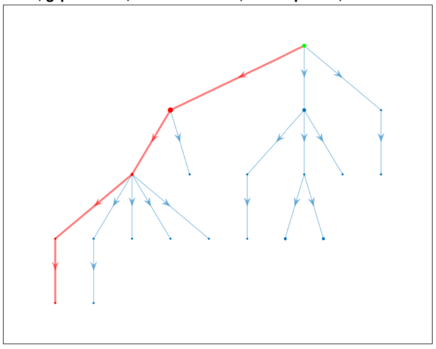

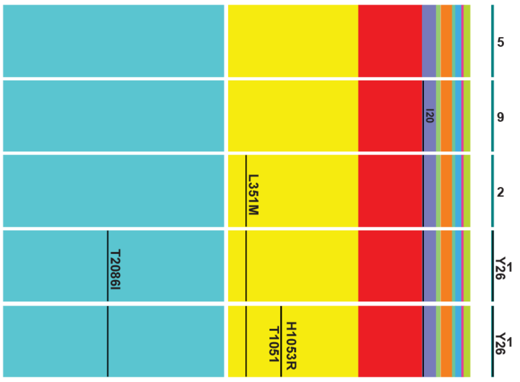

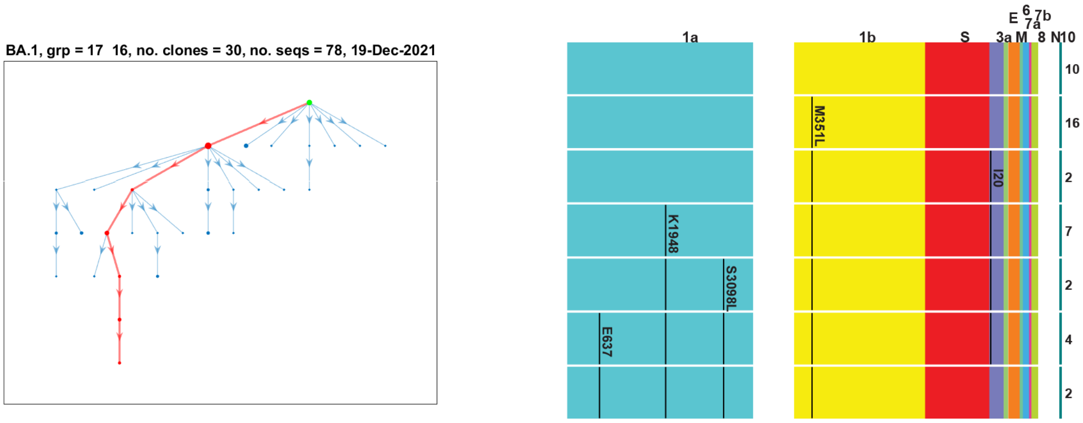
BA.1, grp = 17  16, no. clones = 30, no. seqs = 78, 19-Dec-2021
1a
1b
S
E
6
7b
7a
3a
M
8
N
10
M351L
I20
K1948
S3098L
E637
10
16
2
7
2
4
2

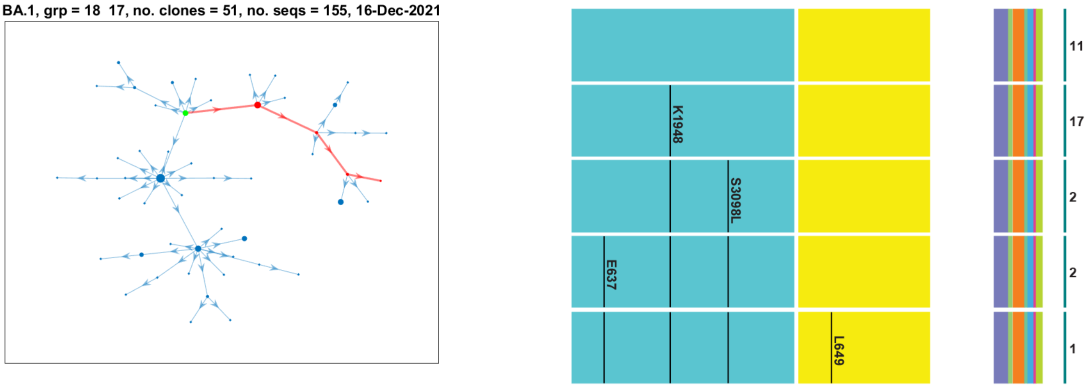
BA.1, grp = 18  17, no. clones = 51, no. seqs = 155, 16-Dec-2021
K1948
S3098L
E637
L649
11
17
2
2
1

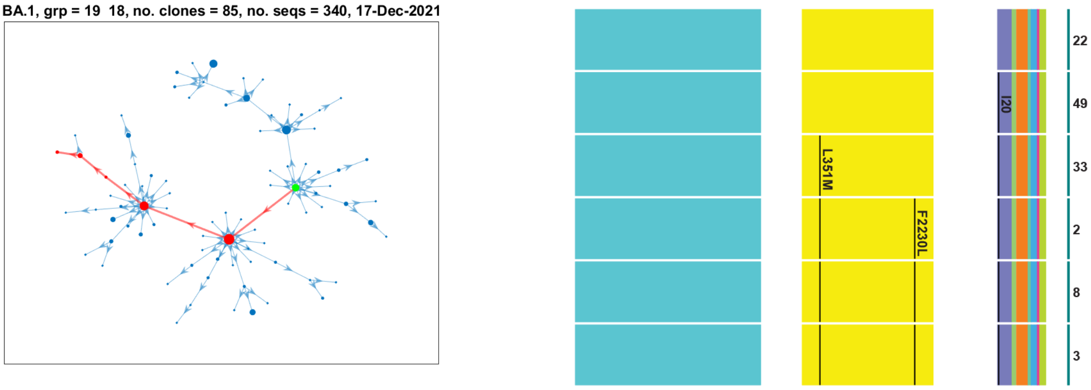
BA.1, grp = 19  18, no. clones = 85, no. seqs = 340, 17-Dec-2021
I20
L351M
F2230L
22
49
33
2
8
3

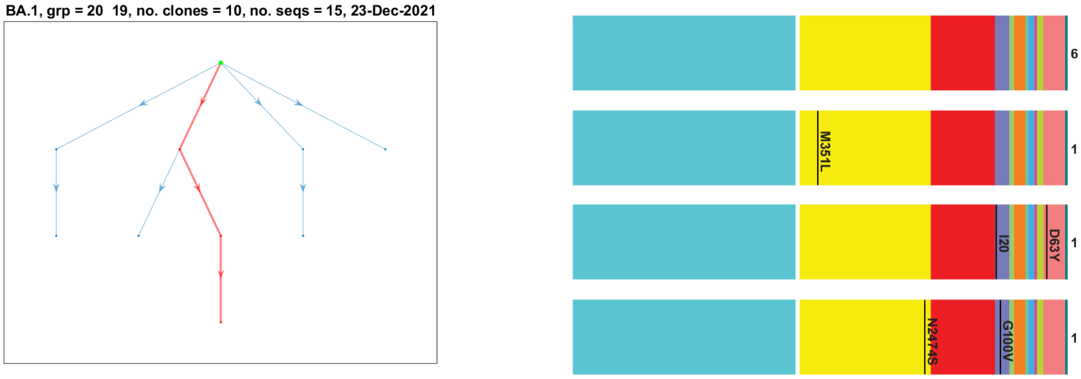
BA.1, grp = 20  19, no. clones = 10, no. seqs = 15, 23-Dec-2021
M351L
I20
D63Y
N2474S
G100V
6
1
1
1

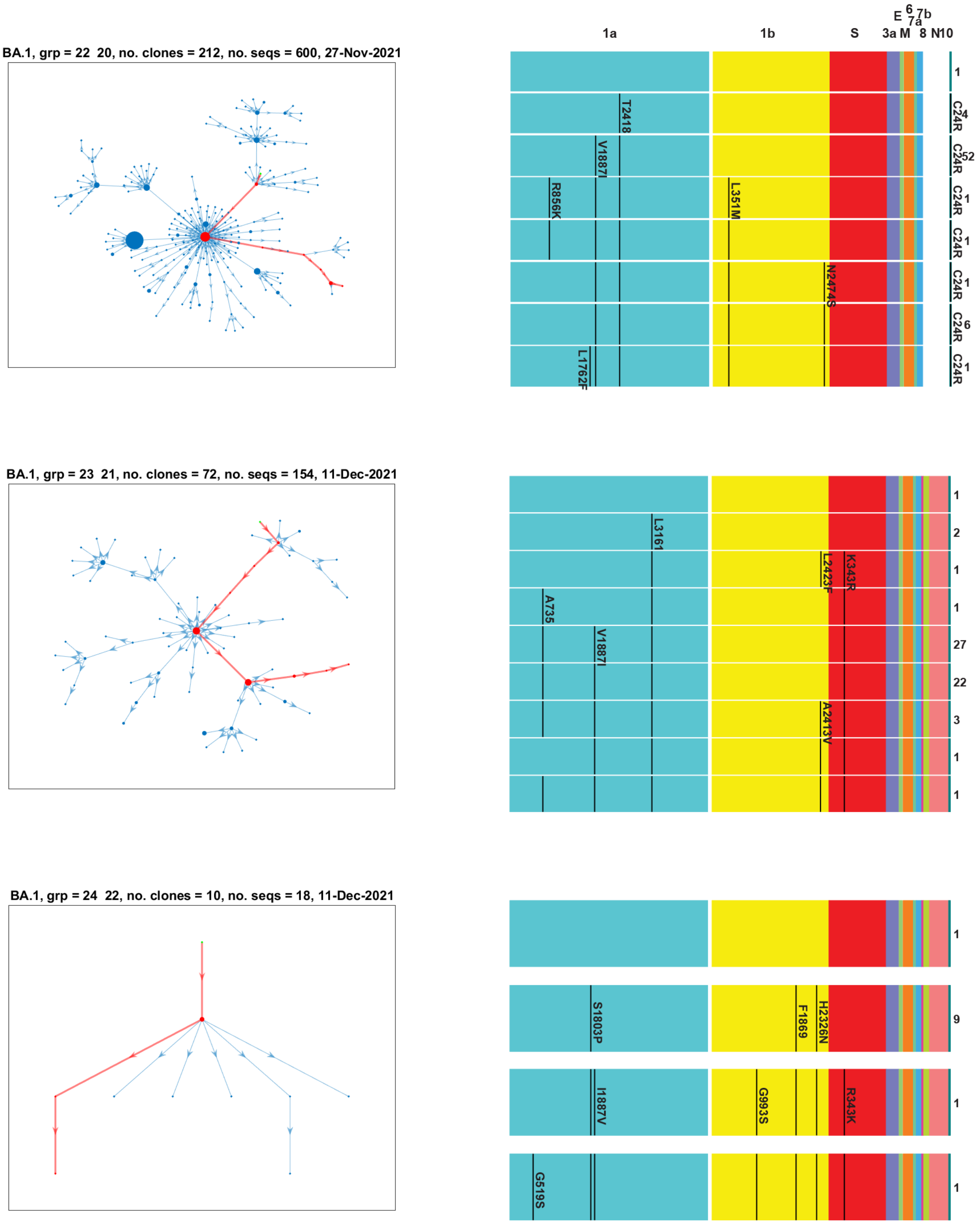


S2 Fig: Estimated clonal transmission networks for all Omicron groups, the longest mutational pathways (red), and the resulting ORF mutations measured between each clone on the path and the root node (green). For groups where the start and stop ORF codons could not be determined, those ORF are depicted as white. The notations above the networks show the lineage, the group numbers within the lineage, the number of clones and sequences included in the network, and the date associated with the root node. The mutational steps for the longest mutational pathway are noted as AA and positions within each ORF (measured from the start of the ORF in that group rather than in terms of the reference sequence), and where these appear in each step as shown by the extent of the black lines. Numbers to the right at each step denote the number of sequences within each clone. A mutation not present in a subsequent step may be due to its loss, or the root node in that clone contains ‘N’s at those positions.

### S3 Fig

#### Power law distributions of clone size

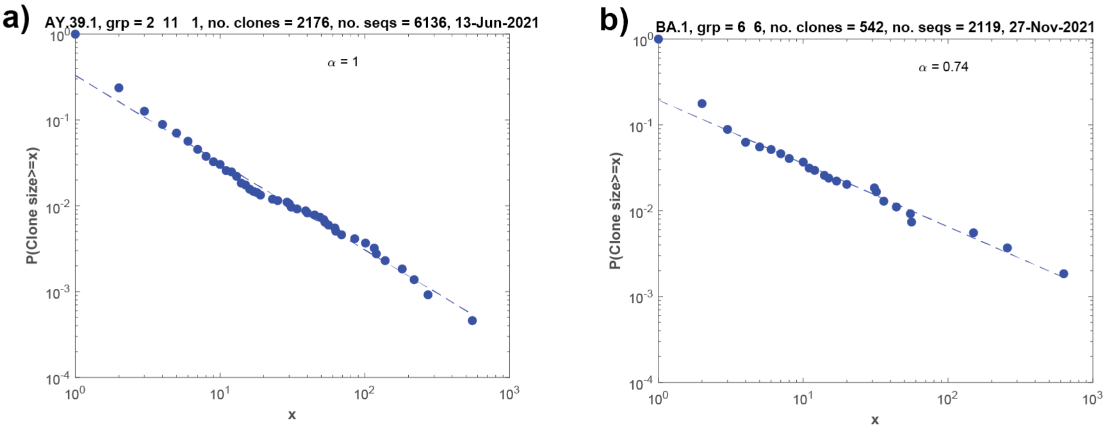


S3 Fig: The fitted power law distributions to clone size for the largest clonal networks of a) Delta, and b) Omicron. Alpha exponents are shown.

### S4 Fig

#### Number of mutations per transmission for Delta

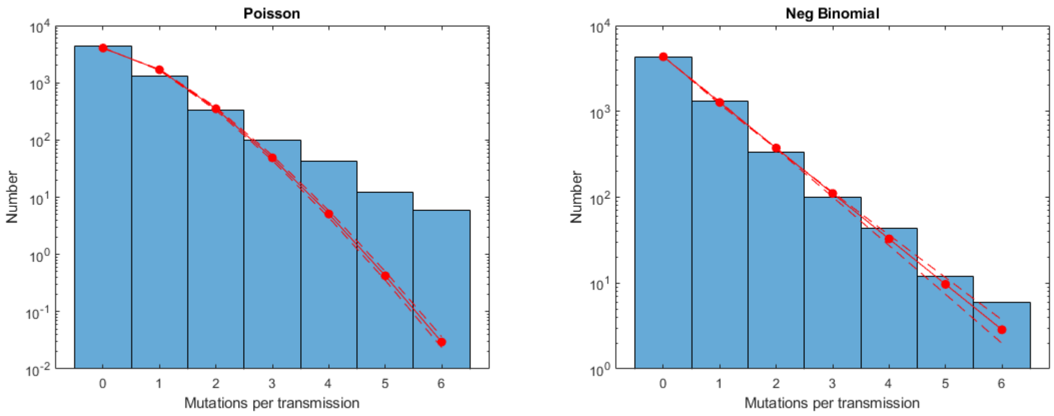


S4 Fig: Fitted a) Poisson ($\lambda = 0.42$), and b) Negative Binomial distributions ($R = 0.99, p = 0.70$) for the number of mutations in each transmission, both between and within clones for the largest Delta clonal network.

### S5 Fig

#### Number of mutations per transmission for Omicron

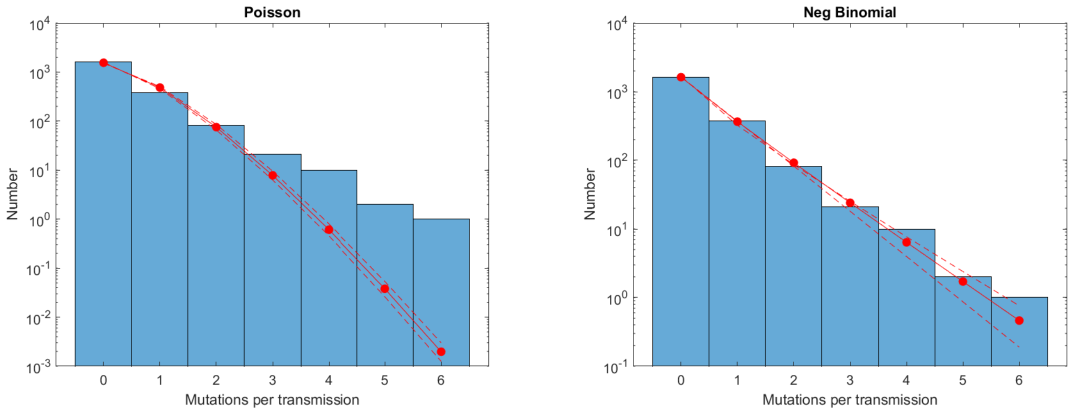


S5 Fig: Fitted a) Poisson ($\lambda = 0.42$), and b) Negative Binomial ($R = 0.80, p = 0.72$) distributions for the number of mutations in each transmission, both between and within clones for the largest Omicron clonal network.

### S1 Table
### Delta mutation probability distributions

| LINEAGE | NO. MUTNS | NO. CLONE EDGES | NO. EDGES IN CLONES | PROB MUTN | ALPHA | $R^2$ | NO. CLONE SIZES | LAMBDA/R | P |
|---|---|---|---|---|---|---|---|---|---|
| **AY.39.1** | 27 | 22 | 15 | 0.73 | 1.66 | 0.97 | 5 | 0.73 | |
| **AY.39.1** | 30 | 18 | 7 | 1.20 | 1.00 | 1.00 | 3 | 1.20 | |
| **AY.39.1** | 25 | 17 | 8 | 1.00 | 2.32 | 1.00 | 4 | 1.00 | |
| **AY.39.1** | 728 | 485 | 421 | 0.80 | 1.12 | 0.94 | 16 | 1.46 | 0.64 |
| **AY.39.1** | 35 | 21 | 15 | 0.97 | 1.32 | 0.99 | 4 | 7.34 | 0.88 |
| **AY.39.1** | 167 | 106 | 91 | 0.85 | 0.97 | 0.82 | 7 | 1.57 | 0.65 |
| **AY.39.1** | 288 | 160 | 103 | 1.10 | 0.97 | 0.93 | 9 | 1.69 | 0.61 |
| **AY.39.1** | 103 | 50 | 13 | 1.63 | 0.83 | 1.00 | 3 | | |
| **AY.39.1** | 31 | 16 | 4 | 1.55 | | | 2 | 12.79 | 0.89 |

| | | | | | | | | | |
|---|---|---|---|---|---|---|---|---|---|
| **AY.39.1** | 29 | 13 | 8 | 1.38 | 1.51 | 1.00 | 4 | 0.83 | 0.38 |
| **AY.39.1** | 2558 | 2175 | 3960 | 0.42 | 1.02 | 0.99 | 44 | 0.99 | 0.70 |
| **AY.39.1** | 313 | 240 | 480 | 0.43 | 0.96 | 0.97 | 17 | 0.97 | 0.69 |
| **AY.39.1** | 583 | 478 | 803 | 0.46 | 1.12 | 0.98 | 22 | 1.10 | 0.71 |
| **AY.39.1** | 347 | 237 | 353 | 0.59 | 0.95 | 0.99 | 15 | 1.06 | 0.64 |
| **AY.39.1** | 75 | 49 | 53 | 0.74 | 1.29 | 0.96 | 8 | 1.41 | 0.66 |
| **B.1.617.2** | 50 | 20 | 12 | 1.56 | 2.96 | 0.98 | 4 | 1.02 | 0.39 |
| **B.1.617.2** | 25 | 11 | 6 | 1.47 | 2.00 | 1.00 | 3 | 0.95 | 0.39 |
| **AY.23** | 44 | 18 | 20 | 1.16 | 1.64 | 0.93 | 4 | 0.50 | 0.30 |
| **AY.39** | 42 | 26 | 8 | 1.24 | 2.32 | 1.00 | 4 | 1.81 | 0.59 |
| **AY.39** | 227 | 165 | 205 | 0.61 | 1.00 | 0.94 | 13 | 1.29 | 0.68 |

| | | | | | | | | | |
|---|---|---|---|---|---|---|---|---|---|
| **AY.39.1.3** | 229 | 109 | 45 | 1.49 | 0.68 | 0.69 | 5 | 1.78 | 0.54 |
| **AY.39.1.3** | 91 | 44 | 17 | 1.49 | 0.78 | 0.91 | 4 | 2.92 | 0.66 |
| **AY.39.1.3** | 603 | 462 | 848 | 0.46 | 0.89 | 0.93 | 19 | 0.89 | 0.66 |
| **AY.39.1.3** | 68 | 48 | 54 | 0.67 | 0.62 | 0.83 | 4 | 2.99 | 0.82 |
| **AY.39.1.2** | 501 | 298 | 93 | 1.28 | 1.13 | 0.83 | 7 | 3.59 | 0.74 |
| **AY.39.1.2** | 200 | 92 | 22 | 1.75 | 0.97 | 0.92 | 4 | 7.46 | 0.81 |
| **AY.39.1.2** | 33 | 18 | 5 | 1.43 | 1.58 | 1.00 | 3 | 5.33 | 0.79 |
| **AY.39.1.2** | 16 | 12 | 20 | 0.50 | 1.09 | 0.91 | 5 | 0.75 | 0.60 |
| **AY.39.1.2** | 1475 | 1154 | 1207 | 0.62 | 1.16 | 0.91 | 22 | 1.34 | 0.68 |
| **AY.39.1.2** | 66 | 39 | 30 | 0.96 | 0.65 | 0.93 | 5 | 8.42 | 0.90 |
| **AY.39.1.1** | 21 | 14 | 46 | 0.35 | 0.22 | 1.00 | 3 | 0.52 | 0.60 |
| | | | **Mean** | **1.00** | **1.22** | | | | |

S1 Table: Probability of mutation per transmission, power exponent of clone size, and distribution of number of mutations per transmission for Delta. If the sample mean exceeded the variance of the number of mutations per transmission, then a Poisson distribution was fitted giving rise to a single reported value of $\lambda$; otherwise the fitted negative binomial distribution was determined and the two parameters $R, p$ are listed.

S2 Table
Omicron mutation probability distributions

| BA.1 GROUPS | NO. MUTNS | NO. CLONE EDGES | NO. EDGES IN CLONES | PROB MUTN | ALPHA | $R^2$ | NO. CLONE SIZES | LAMBDA/R | P |
|---|---|---|---|---|---|---|---|---|---|
| **1** | 31 | 11 | 1 | 2.58 | | | 2 | 2.58 | |
| **2** | 47 | 23 | 4 | 1.74 | | | 2 | 8.15 | 0.82 |
| **3** | 54 | 36 | 23 | 0.92 | 1.41 | 0.81 | 6 | 12.68 | 0.93 |
| **4** | 39 | 24 | 25 | 0.80 | 1.34 | 0.81 | 6 | 1.60 | 0.67 |
| **5** | 6 | 9 | 0 | 0.67 | | | 1 | 1.90 | 0.74 |
| **6** | 33 | 20 | 28 | 0.69 | 1.21 | 0.73 | 6 | 0.90 | 0.57 |
| **7** | 660 | 541 | 1577 | 0.31 | 0.74 | 0.98 | 24 | 0.80 | 0.72 |
| **8** | 43 | 39 | 65 | 0.41 | 1.16 | 0.93 | 9 | 57.97 | 0.99 |
| **9** | 14 | 11 | 7 | 0.78 | 2.32 | 1.00 | 3 | 0.78 | |
| **10** | 112 | 102 | 308 | 0.27 | 0.83 | 0.92 | 17 | 1.14 | 0.81 |
| **11** | 14 | 12 | 12 | 0.58 | 1.55 | 0.94 | 4 | 0.58 | |
| **12** | 16 | 10 | 4 | 1.14 | 1.00 | 1.00 | 3 | 1.15 | 0.50 |
| **13** | 90 | 87 | 68 | 0.58 | 1.26 | 0.91 | 8 | 1.28 | 0.69 |
| **14** | 15 | 11 | 4 | 1.00 | 2.71 | 1.00 | 3 | 3.52 | 0.78 |
| **15** | 14 | 13 | 9 | 0.64 | 0.98 | 0.96 | 4 | 0.64 | |
| **16** | 18 | 19 | 19 | 0.47 | 1.21 | 0.99 | 6 | 0.47 | |
| **17** | 43 | 29 | 48 | 0.56 | 1.17 | 0.99 | 8 | 0.69 | 0.55 |
| **18** | 58 | 50 | 104 | 0.38 | 0.97 | 0.90 | 9 | 0.63 | 0.63 |
| **19** | 105 | 84 | 255 | 0.31 | 0.94 | 0.93 | 15 | 0.46 | 0.60 |
| **20** | 17 | 9 | 5 | 1.21 | | | 2 | 3.02 | 0.71 |

| | | | | | | | | | |
|---|---|---|---|---|---|---|---|---|---|
| **21** | 281 | 211 | 388 | 0.47 | 0.77 | 0.97 | 14 | 0.96 | 0.67 |
| **22** | 98 | 71 | 82 | 0.64 | 0.88 | 0.96 | 9 | 1.12 | 0.64 |
| **23** | 18 | 9 | 8 | 1.06 | | | 2 | 2.21 | 0.68 |
| | | | **Mean** | **0.79** | **1.25** | | | | |

S2 Table: Probability of mutation per transmission, power exponent of clone size, and distribution of number of mutations per transmission for Omicron. If the sample mean exceeded the variance of the number of mutations per transmission, then a Poisson distribution was fitted giving rise to a single reported value of $\lambda$; otherwise the fitted negative binomial distribution was determined and the two parameters $R, p$ are listed.

### S3 Table: GISAID Acknowledgement

We gratefully acknowledge the following authors from the Originating Laboratories responsible for obtaining the specimens and the Submitting

Laboratories where genetic sequence data were generated and shared via the GISAID Initiative, on which this research is based.

| Originating Laboratory | Submitting Laboratory | Authors |
|---|---|---|
| **4Cyte Pathology** | NSW Health Pathology - Institute of Clinical Pathology and Medical Research; Westmead Hospital; University of Sydney | Arnott A., Draper J., Gall M., Martinez E., Rockett R., Sintchenko V., on behalf of ICPMR, CIDM-PH et al |
| **ACT Pathology** | NSW Health Pathology - Institute of Clinical Pathology and Medical Research; Westmead Hospital; University of Sydney | Arnott A., Draper J., Gall M., Martinez E., Rockett R., Sintchenko V., on behalf of ICPMR |
| **Area of Virology, Serology and Virology Division (SAViD), New South Wales Health Pathology Randwick** | Virology Research Laboratory; Area of Virology, Serology and Virology Division (SAViD), New South Wales Health Pathology Randwick | Foster, C.; Au, J.; Ruiz Silva, M.; Deveson, I.; Bull, R.; Van Hal, S.; Rawlinson, W. |
| **Austech Medical Laboratories** | NSW Health Pathology - Institute of Clinical Pathology and Medical Research; Westmead Hospital; University of Sydney | Arnott A., Draper J., Gall M., Martinez E., Rockett R., Sintchenko V., on behalf of ICPMR, CIDM-PH et al |
| **Australian Clinical Labs** | NSW Health Pathology - Institute of Clinical Pathology and Medical Research; Westmead Hospital; University of Sydney | Arnott A., Draper J., Gall M., Martinez E., Rockett R., Sintchenko V., on behalf of ICPMR, CIDM-PH et al. |

| | | |
|---|---|---|
| **Capital Pathology** | NSW Health Pathology - Institute of Clinical Pathology and Medical Research; Westmead Hospital; University of Sydney | Arnott A., Draper J., Gall M., Martinez E., Rockett R., Sintchenko V., on behalf of ICPMR |
| **Capital Pathology** | Schwessinger Lab | Austin Bird, Bayantes Dagvadorj, Scott Ferguson, Robyn Hall, Evie Hodgson, Ashley Jones, Elise Kellett, Karina Kennedy, Rachel Leonard, Sandra Molloy, Carl McCombe, Catalina Barragán Quintero, Rene Riedelbauch, Benjamin Schwessinger, Gabrielle Smith, Paul Whiting, Salome Wilson, Carolina Correa Ospina, Emma Crean, Daniel Yu |
| **Children's Hospital at Westmead** | Viromics lab, Centre for Virus Research, Westmead Institute for Medical Research | John-Sebastian Eden, Rebecca Burrell, Karan Kim, Philip Britton |
| **Childrens Hospital Westmead** | NSW Health Pathology - Institute of Clinical Pathology and Medical Research; Westmead Hospital; University of Sydney | CIDM-PH et al. |
| **Douglass Hanly Moir Pathology** | NSW Health Pathology - Institute of Clinical Pathology and Medical Research; Westmead Hospital; University of Sydney | Arnott A., Draper J., Gall M., Martinez E., Rockett R., Sintchenko V., on behalf of ICPMR, CIDM-PH et al. |

| | | |
|---|---|---|
| **HISTOPATH PATHOLOGY** | NSW Health Pathology - Institute of Clinical Pathology and Medical Research; Westmead Hospital; University of Sydney | Arnott A., Draper J., Gall M., Martinez E., Rockett R., Sintchenko V., on behalf of ICPMR, CIDM-PH et al. |
| **Laverty Pathology** | NSW Health Pathology - Institute of Clinical Pathology and Medical Research; Westmead Hospital; University of Sydney | Arnott A., Draper J., Gall M., Martinez E., Rockett R., Sintchenko V., on behalf of ICPMR, CIDM-PH et al. |
| **Liverpool Hospital** | NSW Health Pathology - Institute of Clinical Pathology and Medical Research; Westmead Hospital; University of Sydney | Arnott A., Draper J., Gall M., Martinez E., Rockett R., Sintchenko V., on behalf of ICPMR, CIDM-PH et al. |
| **Medihealth Pathology** | NSW Health Pathology - Institute of Clinical Pathology and Medical Research; Westmead Hospital; University of Sydney | Arnott A., Draper J., Gall M., Martinez E., Rockett R., Sintchenko V., on behalf of ICPMR |
| **Medlab Pathology** | NSW Health Pathology - Institute of Clinical Pathology and Medical Research; Westmead Hospital; University of Sydney | Arnott A., Draper J., Gall M., Martinez E., Rockett R., Sintchenko V., on behalf of ICPMR, CIDM-PH et al. |

| | | |
|---|---|---|
| **Microbiological Diagnostic Unit** | NSW Health Pathology - Institute of Clinical Pathology and Medical Research; Westmead Hospital; University of Sydney | Arnott A., Draper J., Gall M., Martinez E., Rockett R., Sintchenko V., on behalf of ICPMR |
| **Microbiological Diagnostic Unit - Public Health Laboratory (MDU-PHL)** | Microbiological Diagnostic Unit - Public Health Laboratory (MDU-PHL) | Seemann T., Horan, K., Sait, M.L., Sherry, N.L. |
| **Nepean Hospital** | Viromics lab, Centre for Virus Research, Westmead Institute for Medical Research | John-Sebastian Eden, Rebecca Burrell, Karan Kim, Philip Britton |
| **New South Wales Health Pathology Royal Prince Alfred Hospital** | Microbiology RPAH | Foster, C.; Au, J.; Ruiz Silva, M.; Deveson, I.; Bull, R.; Van Hal, S.; Rawlinson, W. |
| **Other Facility** | NSW Health Pathology - Institute of Clinical Pathology and Medical Research; Westmead Hospital; University of Sydney | Arnott A., Draper J., Gall M., Martinez E., Rockett R., Sintchenko V., on behalf of ICPMR |

| | | |
|---|---|---|
| **PHV-FSS** | PHV-FSS | Chenwei Wang, Liam Mcintyre on behalf of Q-PHIRE Genomics |
| **PathWest** | NSW Health Pathology - Institute of Clinical Pathology and Medical Research; Westmead Hospital; University of Sydney | Arnott A., Draper J., Gall M., Martinez E., Rockett R., Sintchenko V., on behalf of ICPMR |
| **PathWest Laboratory Medicine WA** | PathWest Laboratory Medicine WA Microbial Surveillance UNit | PathWest Laboratory Medicine WA Microbial Surveillance UNit |
| **PathWest Laboratory Medicine WA Microbial Surveillance Unit** | PathWest Laboratory Medicine WA Microbial Surveillance Unit | QEII Medical Centre, Hospital Ave, Nedlands WA 6009 |
| **Pathology North - Central Coast - Wyong Laboratory - NSW Health Pathology** | NSW Health Pathology - Institute of Clinical Pathology and Medical Research; Westmead Hospital; University of Sydney | Arnott A., Draper J., Gall M., Martinez E., Rockett R., Sintchenko V., on behalf of ICPMR |

| | | |
|---|---|---|
| **Pathology North - Gosford Hospital - NSW Health Pathology** | NSW Health Pathology - Institute of Clinical Pathology and Medical Research; Westmead Hospital; University of Sydney | Arnott A., Draper J., Gall M., Martinez E., Rockett R., Sintchenko V., on behalf of ICPMR |
| **Pathology North - Hunter - NSW Health Pathology** | NSW Health Pathology - Institute of Clinical Pathology and Medical Research; Westmead Hospital; University of Sydney | Arnott A., Draper J., Gall M., Martinez E., Rockett R., Sintchenko V., on behalf of ICPMR, CIDM-PH et al. |
| **Pathology North - Hunter New England - Tamworth Laboratory - NSW Health Pathology** | NSW Health Pathology - Institute of Clinical Pathology and Medical Research; Westmead Hospital; University of Sydney | Arnott A., Draper J., Gall M., Martinez E., Rockett R., Sintchenko V., on behalf of ICPMR |
| **Pathology North - Lismore laboratory - NSW Health Pathology** | NSW Health Pathology - Institute of Clinical Pathology and Medical Research; Westmead Hospital; University of Sydney | Arnott A., Draper J., Gall M., Martinez E., Rockett R., Sintchenko V., on behalf of ICPMR |

| | | |
|---|---|---|
| **Pathology North - Mid North Coast - Coffs Harbour Laboratory - NSW Health Pathology** | NSW Health Pathology - Institute of Clinical Pathology and Medical Research; Westmead Hospital; University of Sydney | Arnott A., Draper J., Gall M., Martinez E., Rockett R., Sintchenko V., on behalf of ICPMR |
| **Pathology North - Royal North Shore Hospital - NSW Health Pathology** | NSW Health Pathology - Institute of Clinical Pathology and Medical Research; Westmead Hospital; University of Sydney | Arnott A., Draper J., Gall M., Martinez E., Rockett R., Sintchenko V., on behalf of ICPMR, CIDM-PH et al. |
| **Pathology West - NSW Health Pathology** | NSW Health Pathology - Institute of Clinical Pathology and Medical Research; Westmead Hospital; University of Sydney | Arnott A., Draper J., Gall M., Martinez E., Rockett R., Sintchenko V., on behalf of ICPMR, CIDM-PH et al. |
| **Public Health Virology, Forensic and Scientific Services** | Public Health Virology, Forensic and Scientific Services | Alyssa T. Pyke |

| | | |
|---|---|---|
| **QML Pathology** | NSW Health Pathology - Institute of Clinical Pathology and Medical Research; Westmead Hospital; University of Sydney | Arnott A., Draper J., Gall M., Martinez E., Rockett R., Sintchenko V., on behalf of ICPMR |
| **Queensland Health Forensic and Scientific Services** | Queensland Health Forensic and Scientific Services | Chenwei Wang, Son Nguyen on behalf of Q-PHIRE Genomics |
| **Royal Darwin Hospital Pathology** | Microbiological Diagnostic Unit - Public Health Laboratory (MDU-PHL) | Meumann, E., Baird, R., Caly L., Seemann T., Sait, M.L., Druce J., Horan, K., Sherry, N.L. |
| **Royal Hobart Hospital** | Royal Hobart Hospital | Dr L. Cooley, Mr R Vanhaeften, Christine Guglielmino |
| **SA Pathology** | SA Pathology | Caitlin Selway, Lex Leong, Chuan Kok Lim, Mark Turra, Ivan Bastian, Geoff Higgins |
| **SULLIVAN & NICOLAIDES** | NSW Health Pathology - Institute of Clinical Pathology and Medical Research; Westmead Hospital; University of Sydney | Arnott A., Draper J., Gall M., Martinez E., Rockett R., Sintchenko V., on behalf of ICPMR |
| **Safework Laboratories** | NSW Health Pathology - Institute of Clinical Pathology and Medical Research; Westmead Hospital; University of Sydney | Arnott A., Draper J., Gall M., Martinez E., Rockett R., Sintchenko V., on behalf of ICPMR |

| | | |
|---|---|---|
| **South Eastern Area Laboratory Services (SEALS)** | NSW Health Pathology - Institute of Clinical Pathology and Medical Research; Westmead Hospital; University of Sydney | Arnott A., Draper J., Gall M., Martinez E., Rockett R., Sintchenko V., on behalf of ICPMR, CIDM-PH et al. |
| **Southern. IML Pathology** | NSW Health Pathology - Institute of Clinical Pathology and Medical Research; Westmead Hospital; University of Sydney | Arnott A., Draper J., Gall M., Martinez E., Rockett R., Sintchenko V., on behalf of ICPMR, CIDM-PH et al. |
| **St Vincent's Pathology (SydPath)** | NSW Health Pathology - Institute of Clinical Pathology and Medical Research; Westmead Hospital; University of Sydney | Arnott A., Draper J., Gall M., Martinez E., Rockett R., Sintchenko V., on behalf of ICPMR, CIDM-PH et al. |
| **Sullivan Nicolaides Pathology** | NSW Health Pathology - Institute of Clinical Pathology and Medical Research; Westmead Hospital; University of Sydney | Arnott A., Draper J., Gall M., Martinez E., Rockett R., Sintchenko V., on behalf of ICPMR |
| **Sydney South West Pathology Service (SSWPS) - Concord Repatriation General** | NSW Health Pathology - Institute of Clinical Pathology and Medical Research; Westmead Hospital; University of Sydney | Arnott A., Draper J., Gall M., Martinez E., Rockett R., Sintchenko V., on behalf of ICPMR, CIDM-PH et al. |

| | | |
|---|---|---|
| **Hospital - NSW Health Pathology** | | |
| **Sydney South West Pathology Service (SSWPS) - Liverpool Hospital - NSW Health Pathology** | NSW Health Pathology - Institute of Clinical Pathology and Medical Research; Westmead Hospital; University of Sydney | Arnott A., Draper J., Gall M., Martinez E., Rockett R., Sintchenko V., on behalf of ICPMR, CIDM-PH et al. |
| **Sydney South West Pathology Service (SSWPS) - Royal Prince Alfred Hospital - NSW Health Pathology** | NSW Health Pathology - Institute of Clinical Pathology and Medical Research; Westmead Hospital; University of Sydney | Arnott A., Draper J., Gall M., Martinez E., Rockett R., Sintchenko V., on behalf of ICPMR, CIDM-PH et al. |
| **Territory Pathology** | Territory Pathology | Ella Meumann, Dimitrios Menouhos, Robert Baird |
| **The Children's Hospital at Westmead** | NSW Health Pathology - Institute of Clinical Pathology and Medical Research; Westmead Hospital; University of Sydney | Arnott A., Draper J., Gall M., Martinez E., Rockett R., Sintchenko V., on behalf of ICPMR, CIDM-PH et al. |

| | | |
|---|---|---|
| **Victorian Infectious Diseases Reference Laboratory (VIDRL)** | VIDRL and MDU-PHL | Caly L., Seemann T., Sait, M.L., Druce J., Sherry, N.L. |
| **Virtus Diagnostics - IVFAustralia** | NSW Health Pathology - Institute of Clinical Pathology and Medical Research; Westmead Hospital; University of Sydney | Arnott A., Draper J., Gall M., Martinez E., Rockett R., Sintchenko V., on behalf of ICPMR |
| **unknown** | PHV-FSS | Chenwei Wang, Liam Mcintyre, Son Nguyen on behalf of Q-PHIRE Genomics |
